# Context-specific effects of CCM2 on atherosclerosis

**DOI:** 10.64898/2026.10.27.26366328

**Authors:** Shi Fang, Gavin R. Schnitzler, Amélie Vromman, Jiahao Jiang, Mohita Maurya, Frederick K. Kamanu, James J Park, Ran Cui, Ronodeep Mitra, Allison Gabbert, Cindy Zheng, Lily Widdup, Jathin S. Rao, Natalie Pham, Mengyu Chen, Seid Miad Zandavi, Jesse M. Engreitz, Martin S. Taylor, Nicholas A. Marston, Alec A. Schmaier, Marc S. Sabatine, Peter Libby, Christian T. Ruff, Rajat M. Gupta

## Abstract

Genetic variants in the Cerebral Cavernous Malformation 2 (*CCM2*) locus are associated with coronary artery disease (CAD), but the role of *CCM2* in atherosclerosis is unknown. We previously identified *CCM2* through endothelial cell (EC) Perturb-seq as a regulator of transcriptional programs enriched for CAD risk genes, and predicted that reduced CCM2 function would be athero-protective. Here, we integrated human genetics, functional studies in human aortic ECs, and single cell profiling with mouse models of atherosclerosis to define the effects of reduced CCM2 signaling across disease stages. *Ccm2*^+/-^ mice developed less atherosclerosis after 12 weeks of high-fat, high-cholesterol diet. At baseline, aortic ECs from *Ccm2*^+/-^ mice exhibited reduced inflammatory transcriptional programs, and *CCM2* knockout in human aortic ECs increased *KLF2* expression and reduced monocyte adhesion and procoagulant activity. However, this protective phenotype was not sustained during disease progression. After 12 weeks of high-fat, high-cholesterol diet, *Ccm2*^+/-^ mice exhibited increased circulating TNFα and increased inflammatory programs in aortic ECs and T cells. Consistent with this transition, *Ccm2*^+/-^ showed increased atherosclerotic burden after 20 weeks of atherogenic diet. Human genetic analyses showed a similar context-dependent association. The lead variant in the *CCM2* locus (rs2107732, a missense V53I coding variant in *CCM2*) was associated with lower CAD risk in the general population but higher CAD risk in clinical trial cohorts enriched for cardiometabolic disease. These results demonstrate how the phenotypic consequences of a CAD-associated gene can depend on disease context and stage.

**One Sentence Summary:** CCM2 heterozygosity reduces endothelial inflammation and early atherosclerosis but worsens late-stage disease, consistent with human genetics.

## INTRODUCTION

Coronary artery disease (CAD) is a complex, heritable disease that is the leading cause of death worldwide (*1*). Multiple genome-wide association studies (GWAS) have identified genetic risk loci that account for over 35% of heritability for CAD (*2*). The majority of significant loci are not associated with blood lipids (*3*), and CAD heritability is enriched for enhancers in endothelial and vascular smooth muscle cells (*4*). These observations suggest that vascular function is a central component of CAD risk, and that mechanistic studies of the vascular effects of GWAS risk loci can lead to the development of novel therapies for CAD.

To accelerate the functional characterization of CAD risk variants we recently established a CAD “variant-to-gene-to-program” pipeline using high-throughput CRISPRi-Perturb-seq in endothelial cells (ECs, (*3*)). Using this pipeline, we perturbed all genes within 1 Mb of over 300 CAD GWAS signals and identified five transcriptional programs and 41 candidate genes that were enriched for CAD heritability in ECs. One of the strongest regulators of the CAD-associated programs was cerebral cavernous malformation 2 (CCM2), knockdown of which significantly regulated four out of five of these programs. The *CCM2* locus is significantly associated with risk of CAD in recent GWAS (*2*, *5*). Together with our observations that CCM2 knockdown in ECs increased expression of athero-protective genes, decreased expression of atherogenic genes and improved EC barrier function, this suggested that a quantitative decrease in CCM2 function would protect from the atherosclerosis that underlies CAD. This is in strong contrast to the established effects of complete loss of CCM2 function in brain ECs, which results in the rare, Mendelian disease Cerebral Cavernous Malformations (CCM), characterized by unregulated venous EC proliferation and disrupted endothelial barrier function.

CCM2 acts as a scaffold protein, bridging CCM1/KRIT1 and CCM3/PDCD10 to form the CCM complex (*6–11*). It participates in the regulation of EC barrier function, angiogenesis, and apoptosis (*12*). One key molecular function of the CCM complex is to inhibit MEKK3 kinase activity, which results in downregulation of transcription factors KLF2 and KLF4, normal expression of which is important to maintain healthy EC phenotypes (*9*, *13*). The CCM complex also signals through RhoA & ROCK1/2 to regulate the actin cytoskeleton, and through PI3 kinase to modulate EndMT (the endothelial to mesenchymal transition)(*12*).

Here, we demonstrate the context-dependent role of CCM2 in atherosclerosis using molecular, physiological, and human genetic evidence. *Ccm2*^+/-^ reduced early atherosclerosis in mice (after 12 weeks of atherogenic diet), preceded by a baseline anti-inflammatory transcriptional phenotype in aortic ECs. Concordantly, CCM2 knockout in human aortic ECs showed reduced expression of athero-promoting genes, monocyte adhesion, and procoagulant activity. However, after 12 weeks of atherogenic diet, despite reduced plaque burden, *Ccm2*^+/-^mice showed elevated plasma TNFα and increased inflammatory gene programs in aortic ECs and T cells. The increase of *Klf2* in *Ccm2*^+/-^ ECs at baseline, was also lost at 12 weeks, and this could be recapitulated in CCM2 knockdown human aortic ECs treated with TNFα. This transition to a pro-inflammatory state may underlie the reversed effect in late atherosclerosis, where *Ccm2*^+/-^ mice show increased plaque area at week 20. Human genetic data showed a similar context-dependent association. The likely causal common coding variant in CCM2 was associated with lower CAD risk in the general population but higher risk in clinical trial cohorts enriched for cardiometabolic disease. Taken together, our results indicate that reduced *CCM2* expression protects against the initiation of atherosclerosis, in contrast to the established pathogenic role of strong *CCM2* loss of function in venous malformations in the brain. The discrepancy between early and late stages of atherosclerosis suggests that potential therapeutics to inhibit CCM signaling could be effective, but only to prevent initial atherogenesis or slow early stages of disease.

## RESULTS

### *Ccm2* heterozygous knockout reduces atherosclerotic lesion size in mice

To directly test the effect of CCM2 loss-of-function on atherosclerosis, we studied the effect of heterozygous loss-of-function in a mouse model of hyperlipidemic atherosclerosis. We obtained *Ccm2* heterozygous knockout (*Ccm2*^+/-^) mice generated with a gene-trap insertion in Exon 6 (*14*). These *Ccm2*^+/-^ mice do not develop cerebral cavernous malformations (*14*, *15*). Instead, whole body heterozygous loss-of-function is expected to mimic the partial *CCM2* knockdown we used to connect CCM2 function in ECs to CAD heritability and atherosclerosis-relevant phenotypes (*3*), as well as the global scope of germline variants. Atherosclerosis was induced by a single intravenous injection of AAV2/8 expressing PCSK9 gain-of-function mutation D377Y followed by a 12-week high fat, high cholesterol diet (**Fig. 1A**). The *Ccm2* mRNA levels in the aorta of *Ccm2*^+/-^ mice were significantly lower when compared to *Ccm2^+/+^*mice (**Fig. 1B**). Both genotypes had expected increases in plasma cholesterol in response to AAV2/8-PCSK9-D377Y injection and a 12-week high fat, high cholesterol diet, but there was no significant difference in plasma cholesterol between *Ccm2^+/+^* and *Ccm2*^+/-^ mice (**Fig. 1C**).

**Fig. 1.**
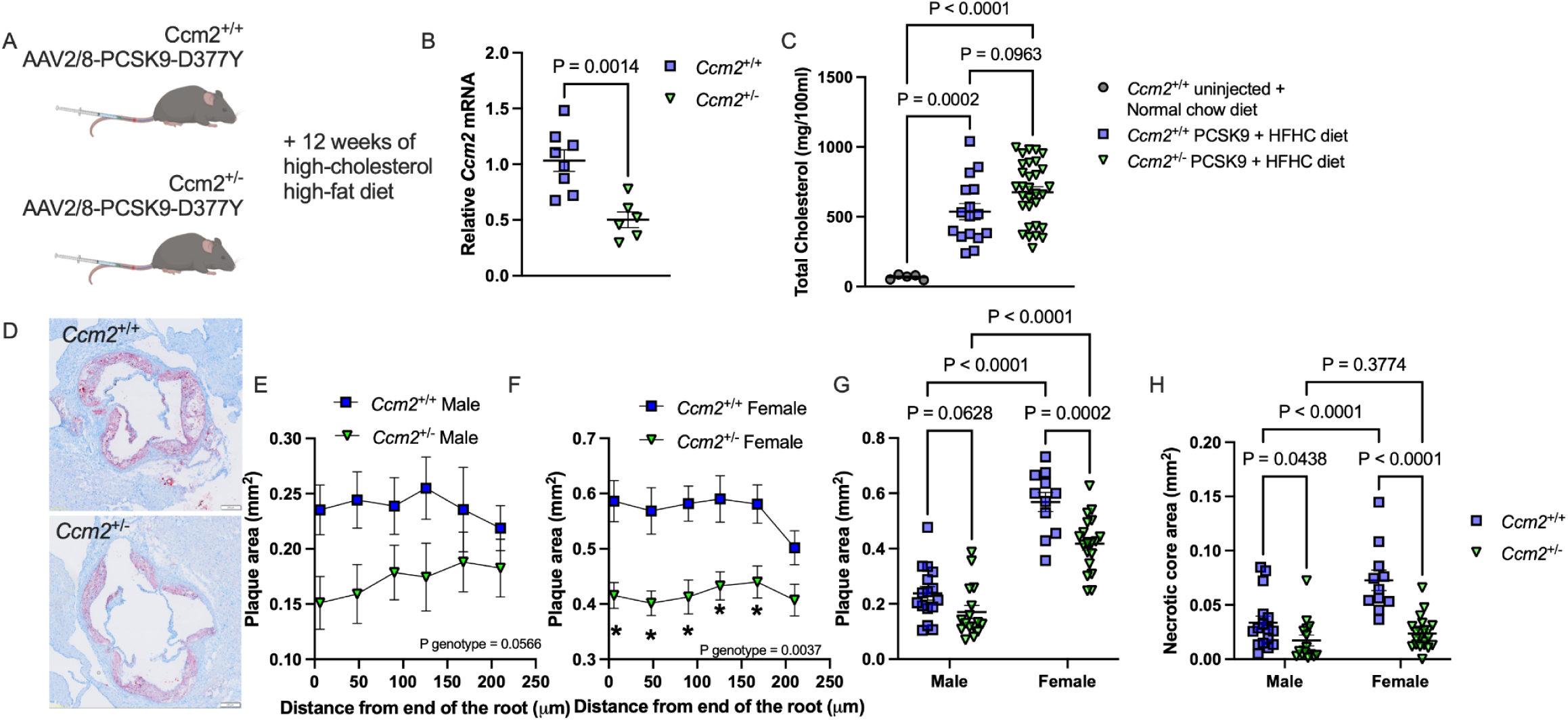
*Ccm2*^+/-^ reduces atherosclerotic lesion size in mice after 12 weeks of atherogenic diet. (**A**) Animal protocol. (**B**) Relative *Ccm2* mRNA levels measured by qPCR in mouse aorta (*Ccm2*^+/+^ n=8; *Ccm2*^+/-^ n=7). (**C**) Total cholesterol levels in plasma at 12 weeks. (**D**) Representative images of atherosclerotic lesions revealed by Oil Red O staining counterstained with hematoxylin at the aortic root from *Ccm2*^+/+^ and *Ccm2*^+/-^ mice. (**E-F**) Atherosclerotic plaque area at each sectioning site every 42 µm along the aortic root in male (**E**; n=13, 14) and female (**F**; n=11, 17) mice. Data are displayed as mean ± SEM. (**G-H**) Average plaque area (**G**) or necrotic core area (**H**) (n= 13, 14, 11, 17, left to right); each dot represents the average plaque or necrotic core area among all six sections from the same mouse. Statistical tests: Unpaired t-test (B), one-way ANOVA with Tukey’s multiple comparisons test (C), two-way ANOVA with repeated measurements (E-F), and two-way ANOVA with Šídák’s multiple comparisons test (G-H).

Average atherosclerotic plaque area at the aortic root was significantly less in *Ccm2*^+/-^mice compared to *Ccm2^+/+^* littermates (**Fig. 1D-G**, 2-way ANOVA P_genotype_ = 0.0024). Female mice developed larger atherosclerotic lesions than male mice (**Fig. 1G**, 2-way ANOVA P_sex_ <0.0001), and previous studies have also shown that female mice with C57BL/6J background have bigger atherosclerotic lesions in AAV-PCSK9 or *Ldlr*^-/-^ models (*16–19*). Although the genotype-associated reduction in lesion area appeared more pronounced in females, the genotype-sex interaction did not reach significance (2-way ANOVA P_interaction_ = 0.1166), indicating the protective effect of *Ccm2*^+/-^ was not statistically distinguishable between sexes. Given this, the fact that the effect of genotype is significant in females but not males may simply be because their greater lesion burden increases the sensitivity to detect the effect of genotype. The effect of *Ccm2* heterozygosity was evident in all the 6 sections that were combined to calculate average plaque area (taken every 42 μm starting from the base of the aortic root) in both male (**Fig. 1E**) and female (**Fig. 1F**) mice. The average of necrotic core area in the atherosclerotic lesions was also lower in *Ccm2*^+/-^ mice when compared to *Ccm2^+/+^* littermates (P_genotype_ <0.0001, **Fig. 1H**). A trend towards smaller lesions was also observed in the aortic arch and brachiocephalic artery in *Ccm2*^+/-^ mice, although there was greater variability, and many mice did not develop lesions at these sites (**fig. S1**). Together these results show a significant reduction in atherosclerosis at 12 weeks of high fat diet in *Ccm2*^+/-^ mice.

### *Ccm2* heterozygous knockout suppresses endothelial inflammatory gene programs in early atherosclerosis

To define the cellular basis of the reduced atherosclerosis in Ccm*2*^+/-^ mice, we performed single-cell RNA sequencing on whole aortas from *Ccm2*^+/-^ and *Ccm2*^+/+^ mice at baseline (Week 0) and with early atherosclerosis (Week 12, **Fig. 2A**). Aortas were dissociated then pooled by genotype and timepoint group (n = 4 per genotype at Week 0, n = 3 at Week 12) and processed for single cell RNA-sequencing. After removal of ambient RNA, doublets and cells with high mitochondrial RNA counts, a total of 51,076 cells were analyzed (**fig. S2A**). Seven aortic cell types were clustered and annotated: ECs, vascular smooth muscle cells (VSMCs), fibroblasts, macrophages, T cells, B cells, and neutrophils (**Fig. 2B, fig. S2B**). In *Ccm2*^+/+^ aortas, *Ccm2* was expressed in all seven cell populations (**fig. S2C**). Among all captured aortic cells at Week 0 or week 12, average *Ccm2* expression was lower in *Ccm2*^+/-^ aortas vs. *Ccm2*^+/+^ (**Fig. 2D**). Concordantly, the expression of *Klf2*, a key EC transcription factor that is normally repressed by Ccm2 signaling, was higher in *Ccm2*^+/-^ aortas (**Fig. 2E**).

**Fig. 2.**
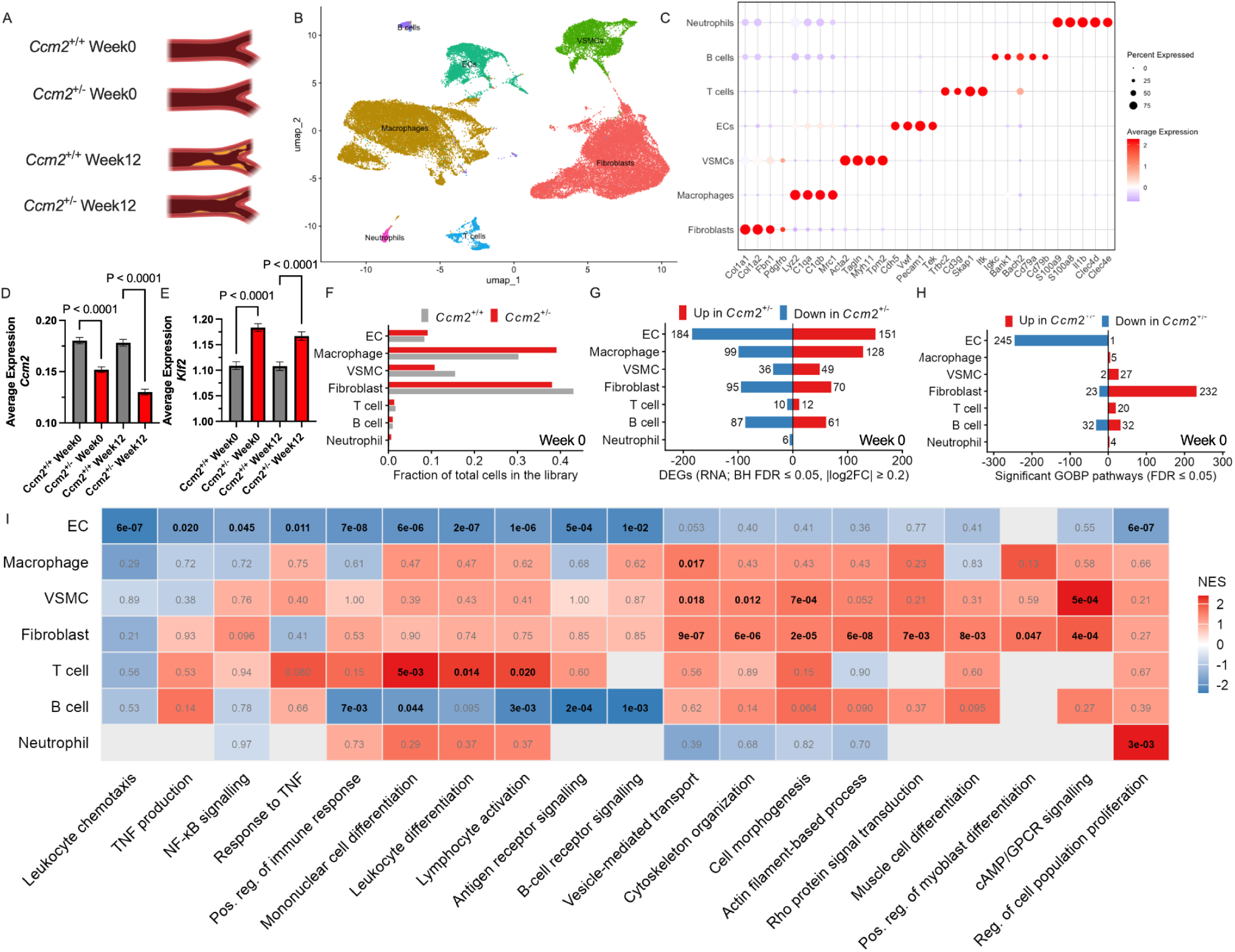
Effects of *Ccm2*^+/-^ on baseline gene expression for each cell type in the aorta. (**A**) scRNAseq study design. (**B**) Major aortic cell types identified by single-cell RNA sequencing. (**C**) Cell type marker gene expression for each cell type. (**D-E**) Average expression of *Ccm2* (**D**) and *Klf2* (**E**) in aortic cells at Week 0, computed with Seurat AverageExpression function. Per cell normalized UMI counts were averaged across all cells in each group. (**F**) Fraction of total cells in the library for each cell type at Week 0. (**G**) Numbers of differentially expressed genes (DEGs) in *Ccm2*^+/-^ versus *Ccm2^+/+^* (*P*adj < 0.05, absolute log_2_FC ≥ 0.2) at Week 0. (**H**) Numbers of GOBP pathways significantly associated (*P*adj <0.05) with all nominally-significant *P*<0.05 DE genes for each cell type at Week 0. (**I**) Normalized enrichment scores (NES) for top cell type-relevant GOBP pathways, ranked by -log_10_FDR, in each aortic cell type. See fig. S3 for the top 5 GO, Reactome and KEGG terms by FDR for each cell type at Week 0. Numbers in boxes are adjusted p.values (grey are *P*adj >0.05).

Focusing on week 0, we found that fibroblasts, macrophages, VSMCs and ECs were the four most abundant cell types for both genotypes (**Fig. 2F**). We performed differential expression analysis on all week 0 cell types (*Ccm2*^+/-^ vs. *Ccm2*^+/+^). ECs and macrophages had the highest number of significant differentially expressed genes (DEGs, 335 and 227 genes at FDR< 0.05, absolute log_2_FC ≥0.2), followed by fibroblasts (165), B cells (148) and VSMCs (85, **Fig. 2G**). To identify the effect of *Ccm2* heterozygosity on gene programs in each cell type, we ran gene set enrichment analysis (GSEA), comparing the differential expression results to Gene Ontology biological process terms (GOBP), as well as KEGG and Reactome terms (**Fig. 2H-I, fig. S3** & **Table S1**). We also examined gene-level effects for genes associated, in the literature, with CAD-relevant cellular functions for the top four vascular cell types (**fig. S4**).

*Ccm2*^+/-^ had the strongest effect on gene programs in ECs (242 FDR<0.05-significant GO terms) and fibroblasts (241), with weaker effects in B-cells (51), VSMCs (32), T-cells (20), macrophages (5) and neutrophils (4, **Fig. 2H**). The most striking pattern was the consistent, significant reduction of inflammatory programs in ECs (**Fig. 2I**), which was also reflected in gene-level reductions of inflammatory transcription factors and cytokines (**fig. S4A**). The most noticeable changes in *Ccm2*^+/-^ macrophages were upregulated efferocytosis genes (e.g. *Mrc1*, *Cd163*, *Gas6*, *Mertk*, and *Timd4*, **fig. S4B**). Fibroblasts showed increases in actin remodeling and muscle cell differentiation, at both the program and gene level, suggesting a shift towards a pro-fibrotic myofibroblast phenotype (**Fig. 2I, fig. S4D**). *Ccm2*^+/-^ VSMCs also showed upregulation in cytoskeletal reorganization programs (**Fig. 2I, fig. S3**) and increases in ECM-remodeling genes (**fig. S4C**), but there was no evidence for a coordinated shift to a synthetic or contractile phenotype or to SMC-foam cells (**fig. S4C**). T cells upregulated differentiation and activation programs (but not cytokine production), B cells downregulated these same programs along with B cell receptor signaling, and neutrophils upregulated a cell proliferation program (**Fig. 2I**).

Together, these findings demonstrate that *Ccm2* heterozygosity produces distinct transcriptional responses, at baseline, across vascular and immune cell populations. Some of these effects, like increased myofibroblast or T cell activation programs, might be expected to have the opposite effect on atherosclerosis than the reduction we observed in *Ccm2*^+/-^ mice at 12 weeks. Instead, the most likely cause of the reduced atherosclerosis in *Ccm2*^+/-^ mice appeared to be the strong, consistent decrease in inflammatory programs and genes in ECs. Since endothelial inflammatory activation and leukocyte recruitment are critical initiating events in atherosclerosis, we focused additional analysis on ECs, while also recognizing potential contributions from other aortic cell types.

To further explore the effects of *Ccm2* heterozygosity on ECs we asked whether the gene expression changes in ECs are driven by a particular endothelial subpopulation. After re-centering and clustering aortic ECs, we identified five subclusters based on their distribution and the distinct expression of subcluster markers (**fig. S5**). Published scRNA-seq studies of the mouse aorta have reported the same five EC subpopulations (*20*, *21*). The three major populations are large vessel luminal ECs (EC1), interstitial/microvascular ECs (EC2), and lymphatic ECs (EC3, **fig. S6**), with EC1 composed of three subclusters: classical arterial ECs (EC1.1), inflammatory ECs with high expression of C1qa and C1qb (EC1.2, (*22*)), and a population with high expression of extracellular matrix remodeling and endothelial-to-mesenchymal transition genes (EC1.3 (*22*)). We performed gene set enrichment analysis (GOBP terms) on DE genes (*Ccm2*^+/-^ vs. *Ccm2*^+/+^) for total ECs and individual EC subclusters. Several GO terms related to cytokine response and production were significantly downregulated in total ECs, and this downregulation was either significant or trending in EC1.1 and EC1.2 (**fig. S5C**). These results suggest that the anti-inflammatory effect in *Ccm2*^+/-^ ECs is primarily associated with the inflammatory subtype EC1.2, with a weaker effect in classical luminal ECs, EC1.1.

### CCM2 loss-of-function suppresses endothelial inflammation and procoagulant in vitro

To validate that the transcriptional effect of CCM2 loss-of-function is associated with athero-protective gene expression, we performed bulk RNA-seq from telomerase-immortalized human aortic endothelial cells (teloHAECs) with either Cas9-mediated homozygous CCM2 knockout (CCM2KO, **fig. S7A**) or CRISPRi-mediated CCM2 knockdown (CCM2KD, **fig. S7B**). In both CCM2KO and CCM2KD teloHAECs, there was a shift toward an athero-protective phenotype. CCM2KO significantly decreased athero-prone genes (*COL4A1*, *COL4A2*, *SELE*, *CCL2*, *EDN1*, *PDGFB* (*3*)) and elevated athero-protective genes (*THBD*, *TNXB*, *NOS3*, *KLF2*) while CCM2KD generally moved these genes in the same directions (**Fig. 3A**).

**Fig. 3.**
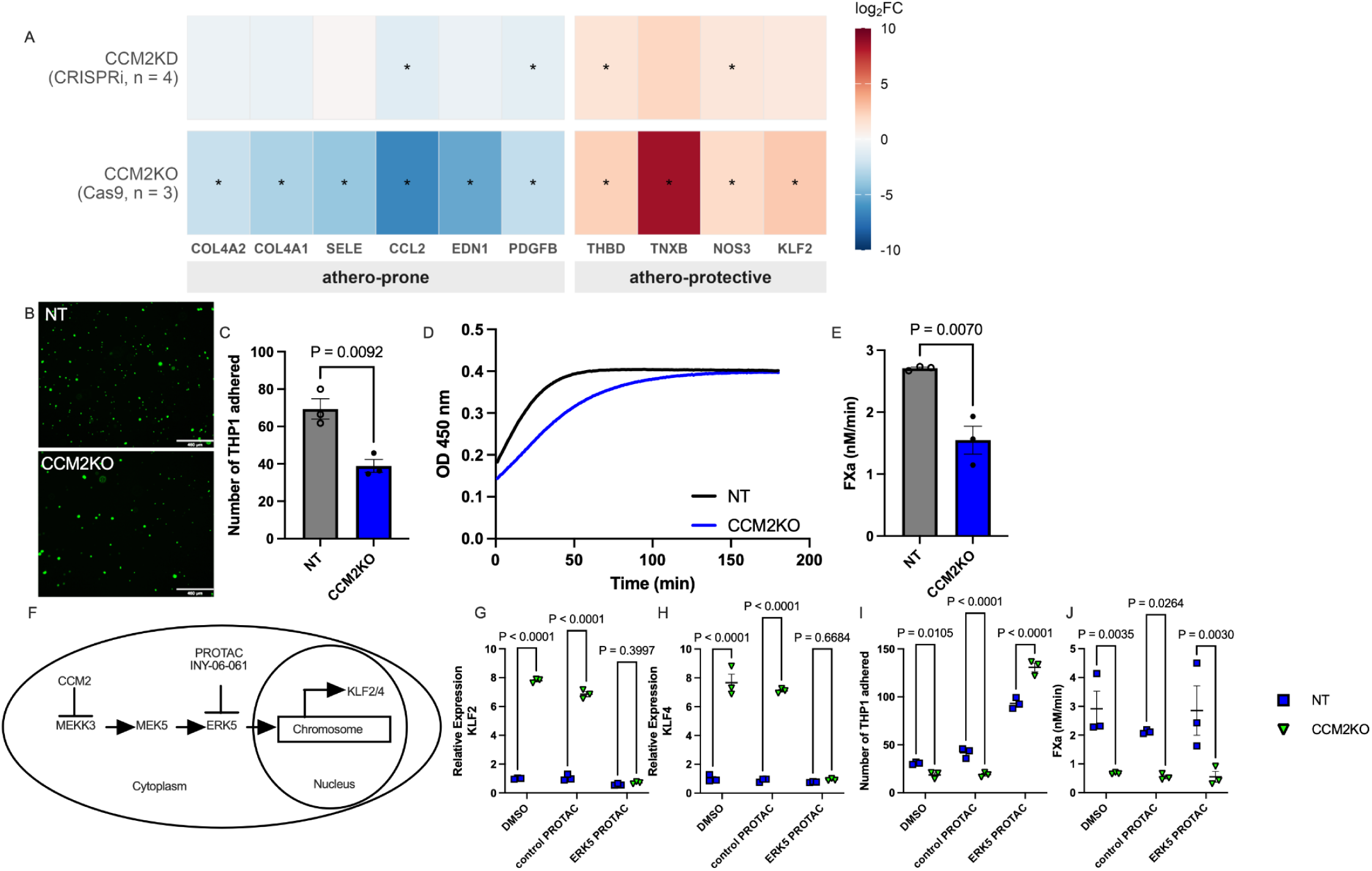
*CCM2* knockout in human ECs supports an anti-inflammatory effect of *CCM2* loss-of-function. **(A)** Heatmap highlighting log2FC changes in athero-promoting or athero-protective gene expression in *CCM2*KO (Cas9, n=3) or *CCM2*KD (CRISPRi, n=4) teloHAECs. *FDR < 0.05. **(B)** Representative images of monocyte adhesion assays in non-targeting (NT) or *CCM2*KO teloHAECs. **(C)** Quantification of monocyte adhesion, as the number of THP1 cells per 1000 sorted TeloHAEC cells. **(D)** Factor X activation kinetics in NT or *CCM2*KO teloHAECs. **(E)** Maximum velocity of Factor X activation (nM/min). **(F)** Diagram of the *CCM2*-MEKK3-ERK5-*KLF2*/*4* signaling axis and the targeted inhibition of ERK5 by PROTAC INY-06-061. **(G**-**H)** Relative mRNA levels of *KLF2* **(G)** and *KLF4* **(H)** in NT or *CCM2*KO teloHAECs treated with DMSO, control PROTAC (INY-06-089), or ERK5 PROTAC (INY-06-061). **(I**-**J)** Monocyte adhesion **(I)** and Factor X activation **(J)** in NT or *CCM2*KO teloHAECs treated with DMSO, control PROTAC (INY-06-089), or ERK5 PROTAC (INY-06-061) (n=3). Bars in (C) & (E): SEM. Each data point represents one biological replicate (biorep), calculated as the average of technical replicates. For (C) & (I) dots are the number of THP-1 cells per 1000 sorted teloHAEC cells from separate wells. Statistical tests: t-test **(C**, **E)** and one-way ANOVA with Tukey’s multiple comparisons test **(G**-**J)**.

Next, we tested the functional outcomes of the anti-inflammatory, athero-protective phenotype observed in the transcriptomics of *Ccm2*^+/-^ mouse aortic ECs and CCM2KO teloHAECs by measuring the adhesion of monocytes to CCM2KO teloHAECs stimulated by TNFα. The number of adhered THP-1 monocytes fell significantly in CCM2KO teloHAECs compared to non-targeting controls (**Fig. 3B-C**). We saw a similar effect in teloHAECs transfected with siRNAs targeting CCM2 (**fig. S7C**). Endothelial inflammatory activation also manifests functionally as increased procoagulant activity (*23*), and we found that CCM2KOs also exhibit slower Factor X activation compared with non-targeting controls (**Fig. 3D-E**).

Previous studies have established that CCM2 loss-of-function activates the MEKK3-ERK5-KLF2/4 signaling pathway (**Fig. 3F**), which is pathological in neurovascular disease (*13*, *24–26*). Consistent with this, CCM2KO cells have greatly increased KLF2 and KLF4 expression (**Fig. 3A, 3G-H “DMSO”**). To test if this axis mediates a protective phenotype in aortic ECs with CCM2 loss-of-function, we treated cells with INY-06-061, a selective proteolysis-targeting chimera (PROTAC) that degrades ERK5, or with an inactive control PROTAC INY-06-089 (**Fig. 3F**) (*27*). KLF2 and KLF4 expression was elevated in CCM2KO cells treated with either DMSO (solvent) or the control PROTAC but returned to the levels in control cells with ERK5 PROTAC (**Fig. 3G-H**). ERK5 PROTAC reversed the reduced monocyte adhesion seen in *CCM2* knockout cells, increasing levels in control cells, but increasing it more in CCM2KO cells (**Fig. 3I**). Concordantly, siRNAs knocking down *KLF2/4* (**fig. S7D-E**) also restored monocyte adhesion in *CCM2* knockout teloHAECs (**fig. S7F**) despite a weaker reduction of *KLF2/4* compared with the ERK5 PROTAC.

The ERK5 PROTAC, however, did not prevent the downregulation of Factor X activation in *CCM2* knockout teloHAECs (**Fig. 3J**), indicating that the effect of CCM2KO on procoagulation may work through an ERK5-KLF2/4-independent pathway. Tissue factor (F3) is the cofactor for Factor VIIa and the two form the extrinsic tenase complex that activates Factor X (**fig. S7G**) (*28*). Because Factor VIIa and Factor X were supplied externally in excess, the rate of Factor Xa generation reflects the relative availability of F3 (which promotes Factor X activation) and tissue factor pathway inhibitor (TFPI, which inhibits Factor X activation, **fig. S7G**). CCM2 knockout reduced F3 expression under every treatment condition including ERK5 degradation (**fig. S7H**). This is consistent with a previous study, using endothelial-specific KLF2/4 double knockout mice, that showed no change in endothelial F3 expression in vivo ((*29*, *30*)). By contrast, the relative expression of TFPI in WT and CCM2KO cells was still altered by PROTAC, indicating some responsiveness to ERK-KLF2 signaling (**fig. S7I**). These observations suggest that the ERK5-independent inhibition of Factor X activation in CCM2KO cells may arise from ERK5-independent suppression of F3 expression.

### CCM2 heterozygosity increases endothelial inflammatory gene programs in established atherosclerosis

Despite the anti-inflammatory state in ECs at baseline, and reduced atherosclerosis at 12 weeks, single cell RNA-seq results from aortas at 12 weeks indicated that the cell-level protective effects do not persist during disease progression. At Week 12, 12,032 cells from *Ccm2*^+/+^ and 11,735 cells from *Ccm2*^+/-^ aortas were collected. Similar to Week 0, the four major cell types were macrophages, fibroblasts, ECs and VSMCs (**Fig. 4A**). While there was some apparent variation in proportions of the 4 major cell types between *Ccm2^+/-^* and ^+/+^ at week 0 (Fig. 2D) and week 12 (Fig. 4A), none of these were found to be significant using the scProportionTest default thresholds (FDR<0.05, fold change > 1.5). At Week 12, ECs had the highest number of DEGs (815 at FDR < 0.05), followed by T cells (168), macrophages (154), fibroblasts (57), VSMCs (41), neutrophils (9) and B cells (0, **Fig. 4B**).

**Fig. 4.**
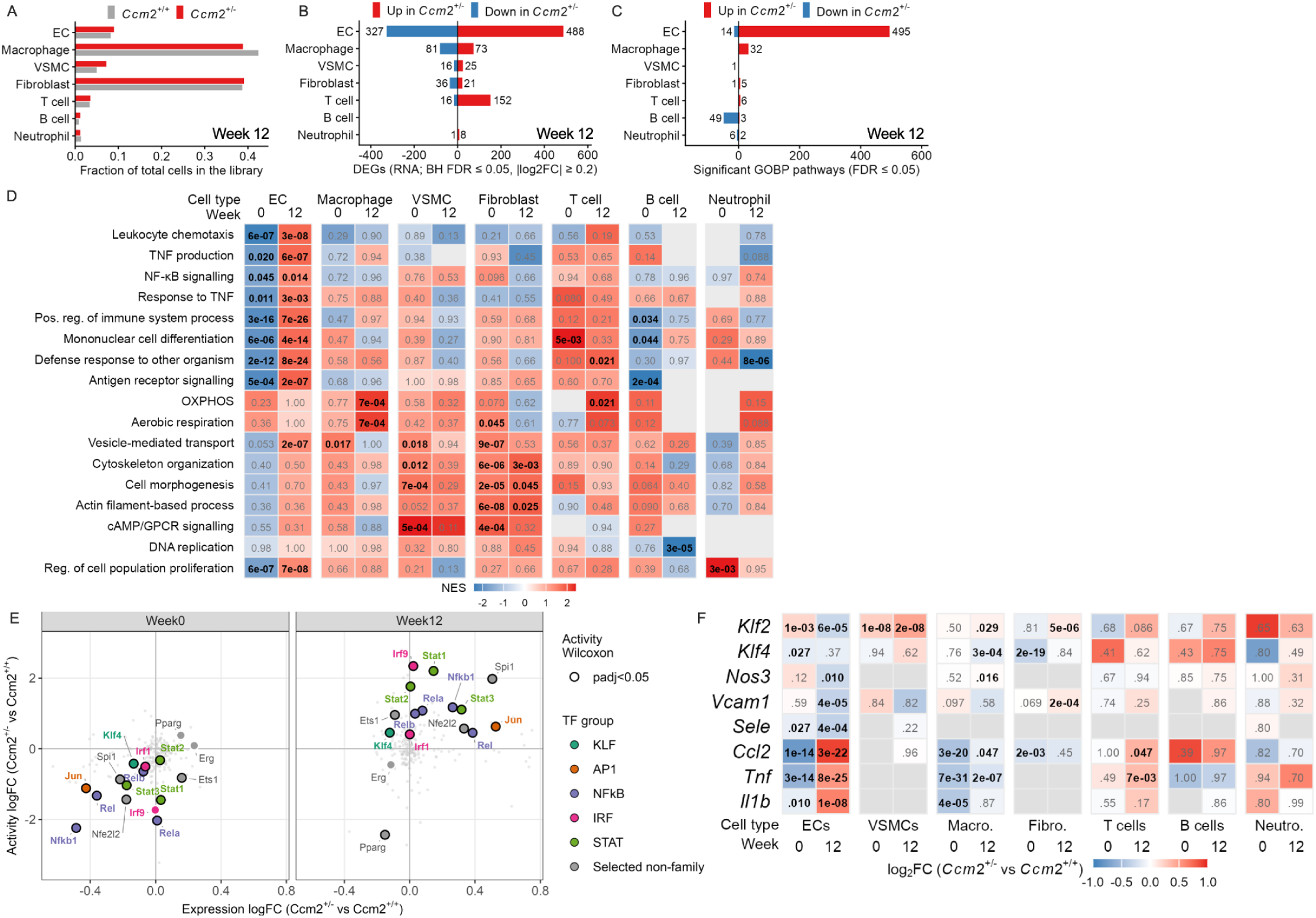
Transcriptional effects of *Ccm2*^+/-^ on each cell type in 12 week atherosclerotic aortas. (**A**) Fraction of total cell counts for each cell type at Week 12. (**B**) Number of differentially expressed genes (P.adj<0.05, |log_2_FC|≥0.2) in each cell type at Week 12. (**C**) Number of GOBP pathways significantly associated (P.adj <0.05) with all nominally significant (p<0.05) DE genes in each cell type at Week 12. See fig. S8 for the top 5 GO, Reactome and KEGG terms strictly by FDR for each cell type. Numbers in boxes are adjusted p values (grey for P.adj > 0.05). (**D**) GSEA normalized enrichment scores (NES) for top cell type-relevant GOBP pathways, ranked by -log_10_FDR, across major aortic cell types at Week 0 and Week 12. (**E**) Inferred activity of transcription factors in the indicated groups/families in aortic ECs at week 0 versus week 12. Dark borders for circles: P.adj < 0.05. (**F**) Log2FC differential expression between *Ccm2*^+/-^ and *Ccm2*^+/+^ of genes for key inflammatory cytokines (*Ccl2*, *Il1b*, *Tnf*), healthy EC functions (*Klf2*, *Klf4*, *Nos3*) and surface adhesion molecules (*Sele*, *Vcam1*) across all cell types and between time points. Numbers in heatmap tiles: FDR.

GO analysis revealed that transcriptional programs changed markedly across cell types with disease progression (**Fig. 4C-D, figs. S8-9, Table S2**). Most notably, inflammatory genes and pathways that were suppressed in *Ccm2*^+/-^ ECs at baseline were significantly increased at Week 12, including inflammatory response, leukocyte chemotaxis, TNF production, and NFκB programs (**Fig. 4D**). This directional reversal was specific to ECs among the major aortic populations examined: 41% of the 536 GO terms in ECs that were significant at either timepoint, were significant at both timepoints but in opposite directions, while this was true for 1% of GO terms for B-cells and 0% for all other cell types. The only other cell type showing a consistent effect of *Ccm2^+/-^* on immune response pathways at week 12 was T cells, which showed significant or trending increases in the same pathways upregulated in ECs (**Fig. 4D**). In addition, macrophages, T cells and neutrophils all showed significant or trending increases in aerobic respiration/oxidative phosphorylation pathways at 12 weeks, suggesting that, in the context of atherosclerosis, *Ccm2^+/-^* amplifies metabolic activity in these cells. Fibroblasts and VSMCs showed significant or trending upregulation of cytoskeletal remodeling programs in *Ccm2^+/-^*aortas at both time points (**Fig. 4D**), suggesting a similar effect on their morphology, migration and/or contractility. At 12 weeks, however, fibroblasts decreased expression of many of the myofibroblast genes that were upregulated at week 0, and ECM production/remodeling genes in fibroblasts and VSMCs largely switched from being increased at week 0 to decreased at week 12 (e.g. *Col1a1*, *Col3a1*, *Col5a1*, **figs. S9C-D**).

The observation that several ECM genes (**figs. S4, S9**) and pathways **(figs. S3, S8)** were upregulated in VSMCs and fibroblasts at baseline but downregulated at week 12 suggested that *Ccm2*^+/-^ might alter the level of fibrosis in week 12 plaques. To test this, we measured aortic root collagen deposition using Masson’s trichrome staining, but found no significant difference between *Ccm2*^+/-^ mice and controls (**fig. S10**).

To assess if the elevated inflammatory gene programs in *Ccm2*^+/-^ ECs at Week 12 reflects a coordinated transcriptional reprogramming, we utilized decoupleR (*31*) with DoRothEA regulons to infer transcription factor (TF) activity in aortic ECs at both time points. TFs of the pro-inflammatory TF families, NF-κB (*Nfkb1*, *Rela*), STAT (*Stat1*, *Stat3*), and IRF (*Irf1*, *Irf9*), were suppressed in *Ccm2*^+/-^ ECs at Week 0 but became more active at Week 12 (**Fig. 4E**), suggesting that changes in inflammatory TF activities drive the observed changes in DE genes.

To further illustrate the transcriptional changes between weeks 0 and 12, we compared the effects of *Ccm2*^+/-^ on the expression of key athero-protective or atherogenic/inflammatory genes across cell types and at both time points (*32–35*) (**Fig. 4F**). This showed, for instance, that the inflammatory cytokines *Ccl2*, *Tnf* & *Il1b*, which decreased significantly in ECs at week 0, are increased in ECs as well as T cells at week 12 and are less strongly decreased in macrophages. For ECs, however, this is not simply a switch to an overall pro-inflammatory state, since the adhesion molecules *Vcam1* and *Sele* (that mediate immune cell recruitment) are more strongly reduced in ECs at 12 weeks than at 0 weeks. Interestingly, *Klf2* expression, which was significantly upregulated in *Ccm2*^+/-^ ECs at Week 0, was significantly reduced at Week 12 (**Fig. 4F**). By contrast, all other major cell types showed significant or trending increases in *Klf2* expression at 12 weeks, consistent with the expected effect of *Ccm2* knockdown on the CCM-MEKK3-KLF2 signaling pathway.

Analysis of DE genes in week 12 EC subpopulations showed that the EndMT-like EC1.3 subtype had the strongest enrichment for cytokine-related GO terms, with lesser but significant enrichment in immune cell-like EC1.2 (**fig. S11A, fig. S12**). The proportions of EC subtypes for each genotype differed between 0 and 12 weeks. In *Ccm2*^+/+^ aortas EC1.3 and EC2 increase by over 1.5-fold, but in *Ccm2*^+/-^ EC1.1 and EC2 decrease, while EC1.2, EC1.3 and EC3 increase (**fig. S11B-C**, (*36*)). To explore the propensity of week 0 ECs subtypes to be converted to (or replaced by) other EC subtypes at week 12, we used CellRank2. The results suggest that all week 0 subtypes are less likely to be EC1.1 at week 12 in *Ccm2*^+/-^, and more likely to be EC1.2 or EC1.3 (**fig. S11D-E**). CellRank2 only infers the most likely status of cells from Week 0 as atherosclerosis progresses, and does not demonstrate lineage transitions. Nonetheless, these results suggest that, as atherosclerosis develops, the *Ccm2*^+/-^ genotype promotes the formation of inflammatory EC1.2 and EndMT-associated EC1.3 at the expense of canonical EC1.1 cells.

Taken together, the response of vascular cells to reduced CCM2 signaling was not uniform, but consistent with a general shift from an anti-inflammatory baseline state to a nuanced, but overall pro-inflammatory state during the development of atherosclerosis. The most striking effect is the switch, in ECs, from decreased cytokine production to increased, which could potentially be connected to the non-canonical decrease of *Klf2* expression observed specifically in 12 week *Ccm2*^+/-^ ECs.

### *Ccm2* heterozygosity increases late-stage atherosclerosis, consistent with increased localized and systemic inflammation in early-stage atherosclerosis

To examine why *Ccm2*^+/-^ ECs switch from anti-inflammatory baseline state to a pro-inflammatory state in 12 week atherosclerosis, we measured circulating cytokines. We found that plasma TNFα increased significantly in *Ccm2*^+/-^ mice at Week 12 (**Fig. 5A**), together with trends towards increases in IL-12p70 and IL-2 (**fig. S13A-B**). The anti-inflammatory cytokine IL10 was also elevated, likely reflecting a compensatory response to the heightened inflammatory environment (consistent with other reports (*37*, *38*)**, fig. S13C**).

**Fig. 5.**
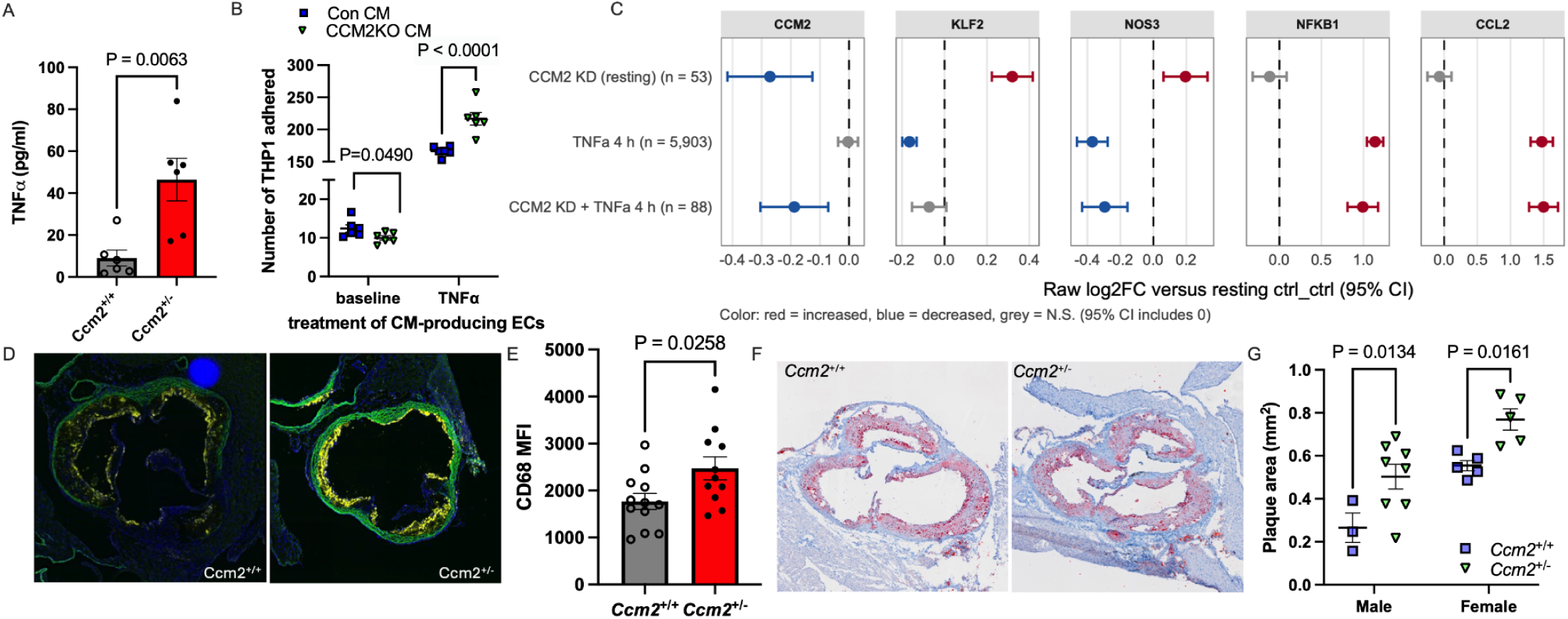
Inflammatory changes in *Ccm2*^+/-^ 12 week mice may underlie a transition to increased atherosclerosis at 20 weeks. (A) Plasma TNFα levels at Week 12 (male and female). (**B**) Monocyte adhesion to teloHAECs incubated with conditioned media (CM) from control or *CCM2*KO teloHAECs. The CM-producing ECs were unstimulated (baseline) or TNFα-stimulated. Dots are the number of THP-1 cells per 1000 sorted TeloHAEC cells from separate wells. (**C**) Expression of *CCM2*, *KLF2*, *NOS3*, *NFKB1*, and *CCL2* in CRISPRi-mediated *CCM2*KD teloHAECs (unstimulated, TNFα-stimulated, and *CCM2*KD + TNFα). (**D-E**) CD68 immunofluorescent staining in aortic roots at Week 12; representative images (**D**) and quantification (**E**, male and female). (**F-G**) Oil Red O staining of atherosclerotic lesions in the aortic root at Week 20; representative images (**F**) and quantification (**G**). Data are mean ± SEM. Statistical tests: Student’s t-test (**A,E**), two-way ANOVA (**B, G**).

Given the elevated circulating TNFα and aortic endothelial inflammatory programs in *Ccm2*^+/-^ mice at Week 12, we next asked whether CCM2 deficiency modulates the production of soluble inflammatory mediators. We performed monocyte adhesion assays on control teloHAECs treated with conditioned media (CM) collected from CCM2KO or control teloHAECs. CM from TNFα-treated CCM2KO teloHAECs led to more monocyte adhesion than CM from TNFα-treated control teloHAECs, whereas CM from baseline/unstimulated CCM2KO teloHAECs resulted in decreased monocyte adhesion (**Fig. 5B**). This suggests that the increased circulating TNFα in *Ccm2^+/-^* mice at 12 weeks could contribute to a functionally-relevant increase in cytokine gene expression in aortic ECs. Conversely, when control and CCM2KO ECs received conditioned medium derived from control ECs, CCM2KO had lower monocyte adhesion regardless of the TNFα treatment status of CM donor cells (**fig. S14**). This is consistent with the persistent suppression of adhesion molecules in *Ccm2^+/-^* ECs at 12 weeks *in vivo* (**Fig. 4F**), and may explain why CCM2KO ECs directly treated with TNFα had lower THP-1 binding than controls (**Fig. 3 B-C**).

To examine this context-dependent effect of *CCM2* loss-of-function in human aortic ECs on a transcriptional level, we performed single cell Perturb-seq experiments in teloHAECs expressing dCas9-KRAB (*3*). As expected, in the absence of TNFα CRISPRi-mediated *CCM2* knockdown resulted in decreased *CCM2* expression while increasing the levels of *KLF2* and its target gene *NOS3*. These increases in *KLF2* and *NOS3*, however, were lost in CCM2KD cells stimulated with TNFα **(Fig. 5C**). CCM2KD did not greatly alter basic EC responses to TNFα, since the TNFα responsive genes *CCL2* and *NFKB1* were increased in both control and knockdown cells. These observations agree with the mouse aortic scRNAseq analysis of ECs, and support a model where some aspects of the protective effect derived from CCM2 loss-of-function (especially those related to KLF2 and its downstream targets) can be abolished in the pro-inflammatory context of established atherosclerosis.

We performed aortic root immunofluorescent staining for the macrophage marker CD68 at Week 12. Aortic roots from *Ccm2*^+/-^ mice showed significantly higher mean fluorescence intensity compared with *Ccm2*^+/+^ littermates (**Fig. 5D-E**). This suggests that, despite reduced adhesion molecules on *Ccm2^+/-^*ECs at 12 weeks, the heightened inflammatory environment (increased circulating TNFα and cytokine production from ECs) combines to promote transmigration, retention, survival and/or proliferation of macrophages in the plaque.

These observations suggest that increased inflammation in early atherosclerosis (12 week) may cause the protective effect of Ccm2 heterozygosity to be reduced or even reversed in late stage atherosclerosis. Indeed, *Ccm2*^+/-^ mice showed significantly increased atherosclerotic plaque area in the PCSK9-D377Y model after 20 weeks of high fat diet (**Fig. 5F-G, fig. S15**).

In summary, these observations show that the anti-inflammatory effects of *Ccm2*^+/-^ found at baseline/Week 0 were not sustained as atherosclerosis developed. Instead, *Ccm2*^+/-^ ECs exhibited higher inflammatory gene programs at Week 12, which may be driven in part by EC responses to increased circulating TNFα. Together with increased accumulation of macrophages in week 12 *Ccm2*^+/-^ plaques, this appears to tip the balance so that *Ccm2*^+/-^ switches from being protective for early stage atherosclerosis (12 week) to being detrimental for late stage atherosclerosis (20 week).

### The CAD-protective association of the CCM2 V53I variant is not sustained in higher-risk cardiovascular clinical trials

The two largest multi-ancestry GWASs for CAD identified a locus of association in the chromosome 7p13 locus (*2*, *5*). We conducted a focused analysis of this region to identify the likely causal variant(s). In the CARDIoGRAMplusC4D meta-analysis of 1.3 million participants, which is the largest published CAD GWAS, we found the two most significant variants in the *Chr7p13* locus are a missense variant in exon 2 of the CCM2 gene (**Fig. 6A**, lead variant: rs2107732, odds ratio = 0.952, *P*= 2×10^-8^, MAF = 0.06) and a non-coding variant (rs73105827, MAF= 0.03) that is 3kb upstream of the *CCM2* promoter. Both variants are in close linkage disequilibrium (LD) with each other (r^2^ = 0.94 among European populations (*39*), **fig. S16A**). Both variants have significant associations with CAD and with diastolic blood pressure (DBP, **Fig. 6B, fig. S16B**), and both showed a strong colocalization signal between the DBP GWAS (GCST90310295) and the CAD GWAS (GCST90132314, PP.H4 = 0.9974, **fig. S16C-E**). The lead variants for both traits are the same, rs2107732 and rs73105827, with the highest colocalization metrics (PP.H4 = 0.521 and 0.464, respectively). Whole-blood eQTL signals for CCM2 at this locus did not colocalize with the CAD association (PP.H4 = 0.299, **fig. S16F-G**). However, causal variant(s) in the locus may affect CCM2 protein activity or exert a regulatory effect in specific cell types or conditions that are not reflected in whole blood.

**Fig. 6.**
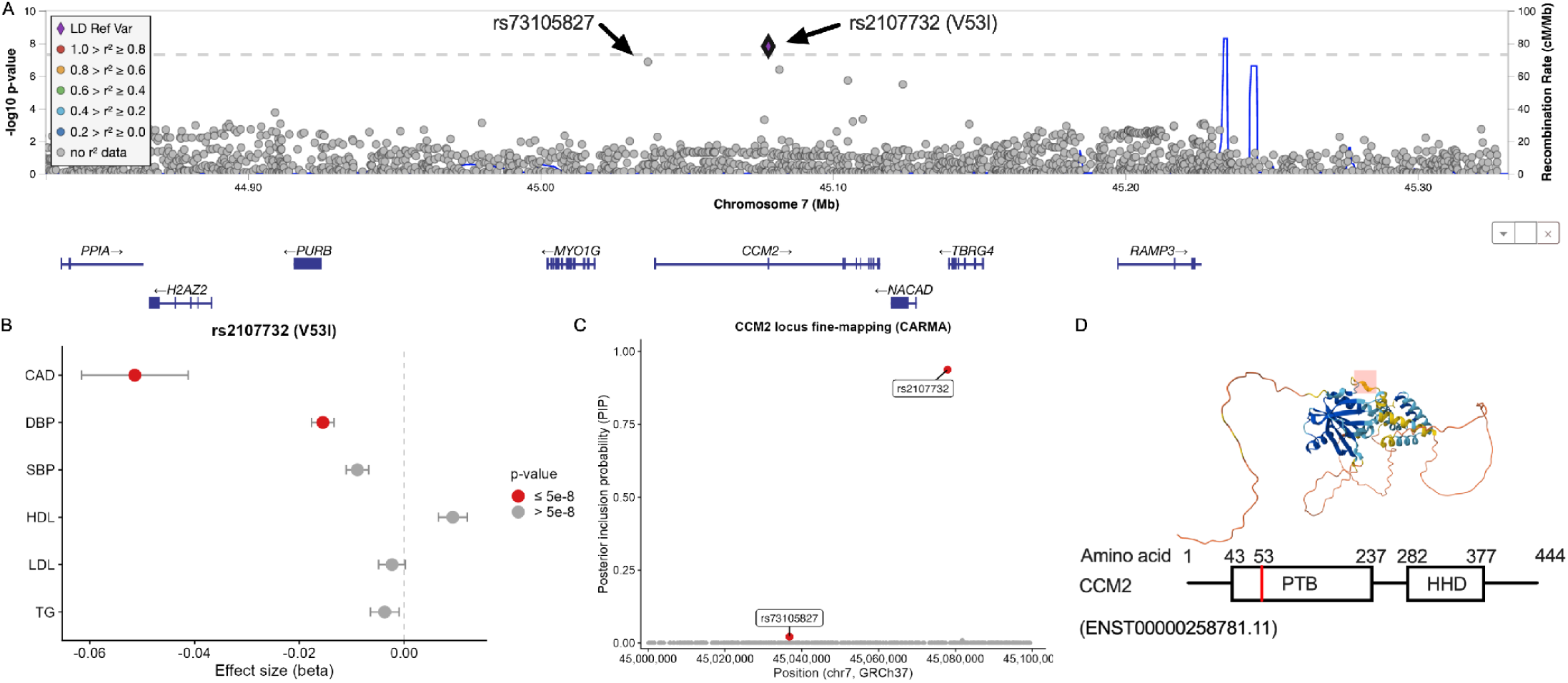
*CCM2* coding variant rs2107732 (V53I) is prioritized as the likely causal variant associated with decreased CAD risk at *Chr7p13*. (A) LocusZoom plot of the *Chr7p13* locus showing −log10 *P* values for variants surrounding *CCM2* from a recent CAD GWAS. (B) PheWAS of rs2107732 demonstrating its association with decreased risk of coronary artery disease (CAD) and diastolic blood pressure (DBP). Red indicates associations reaching genome-wide significance (*P* ≤ 5.00 × 10^-8^). CAD, coronary artery disease; DBP, diastolic blood pressure; SBP, systolic blood pressure; HDL, high-density lipoprotein; LDL, low-density lipoprotein; TG, triglyceride. (C) Posterior inclusion probabilities (PIPs) for variants within 1 Mb of *CCM2*, calculated using the Bayesian fine-mapping algorithm CARMA. (D) Location of rs2107732 within the AlphaFold-predicted *CCM2* protein structure and *CCM2* domain architecture. The variant encodes a valine-to-isoleucine substitution at amino acid 53 within the phosphotyrosine-binding (PTB) domain. V53I is highlighted in red. PTB, phosphotyrosine-binding domain; HHD, harmonin homology domain.

To better identify the lead variant(s) in this locus, we applied three independent Bayesian fine-mapping algorithms (CARMA (*40*), SuSiE (*41*) and FINEMAP (*42*)), along with a recent modification of SuSiE which takes into account the polygenic effects for multiple variants in the locus (SuSiE-inf (*43*)), to disease-association data from the CAD GWAS meta-analyses. All four approaches identify the CCM2 coding variant as the highest-confidence causal variant, with posterior inclusion probabilities (PIP) between 0.73 and 0.94, out of a maximum probability of 1.0 (**Fig. 6C, fig. S16H**). The second highest likelihood causal variant is the noncoding rs73105827, with a maximum PIP of 0.11 (with the SuSiE-inf method). To test if the rs73105827 variant might regulate CCM2 transcription as part of a functional enhancer, we deleted 116bp centered on this variant in teloHAECs, but found that this deletion did not alter the level of *CCM2* mRNA (**fig. S16I**). Hence, we prioritized the missense variant rs2107732, which converts Val53 to Ile in the phosphotyrosine binding domain of CCM2, as the most likely causal variant in this locus (**Fig. 6C-D, fig. S16J**).

To determine whether the context-dependent effects of *Ccm2*^+/-^ mice in atherosclerosis are reflected in humans, we examined the lead CCM2 common missense variant V53I (rs2107732, G>A) across distinct cardiovascular populations using logistic regression. In UKBiobank, in which subjects were not pre-selected for having CAD or CAD risk factors, the minor A allele confers lower odds of CAD (**Fig. 7A, top**). Compared with GG individuals, CAD odds were lower in AG heterozygotes and further reduced in AA homozygotes (**fig. S17A**), indicating an allele-dose-dependent pattern. By contrast, among 68,808 patients with cardiometabolic diseases from six clinical trial cohorts (referred to here as the TIMI trial meta-analysis, see Table 1 and the Methods section for details, (*44–49*)), V53I carriers had higher odds of CAD (incident and prevalent combined, OR 1.108, 95% CI 1.057-1.162, P<0.001, **Fig. 7A, bottom**). With both analyses adjusted for age, sex, and the first 5 principal components of population structure, the association differed significantly between the two populations (ratio of ORs 1.195, 95% CI 1.128-1.266, P_interaction_ = 1.34×10^-9^).

**Fig. 7.**
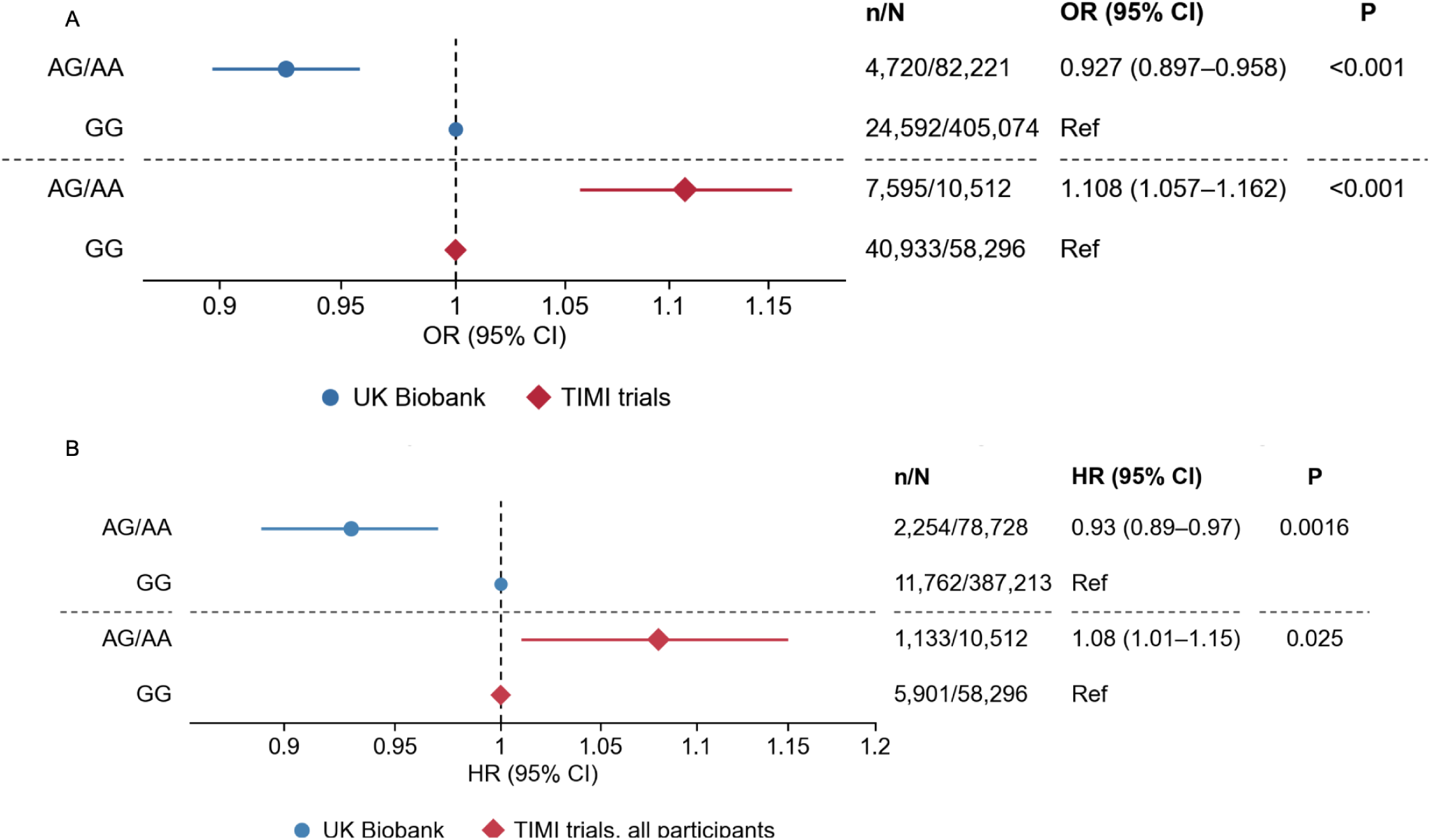
The *CCM2* coding variant rs2107732 (V53I) exhibits context-dependent effects on CAD risk. (**A**) Association of the rs2107732 minor allele (A) with coronary artery disease (incident and prevalent) in the general population (UK Biobank, top) and in patients with cardiometabolic diseases (TIMI trial meta-analysis, bottom). Logistic regression was used. (**B**) Association of rs2107732 with incident CAD (CAD events occurring after enrollment) in UK Biobank participants without CAD at baseline (top) and subsequent coronary events (revascularization/MI/CHD death) among the TIMI trial participants (bottom). Cox regression was used. n/N: CAD cases (n) among all participants (N) of the indicated genotype.

**Table 1.**
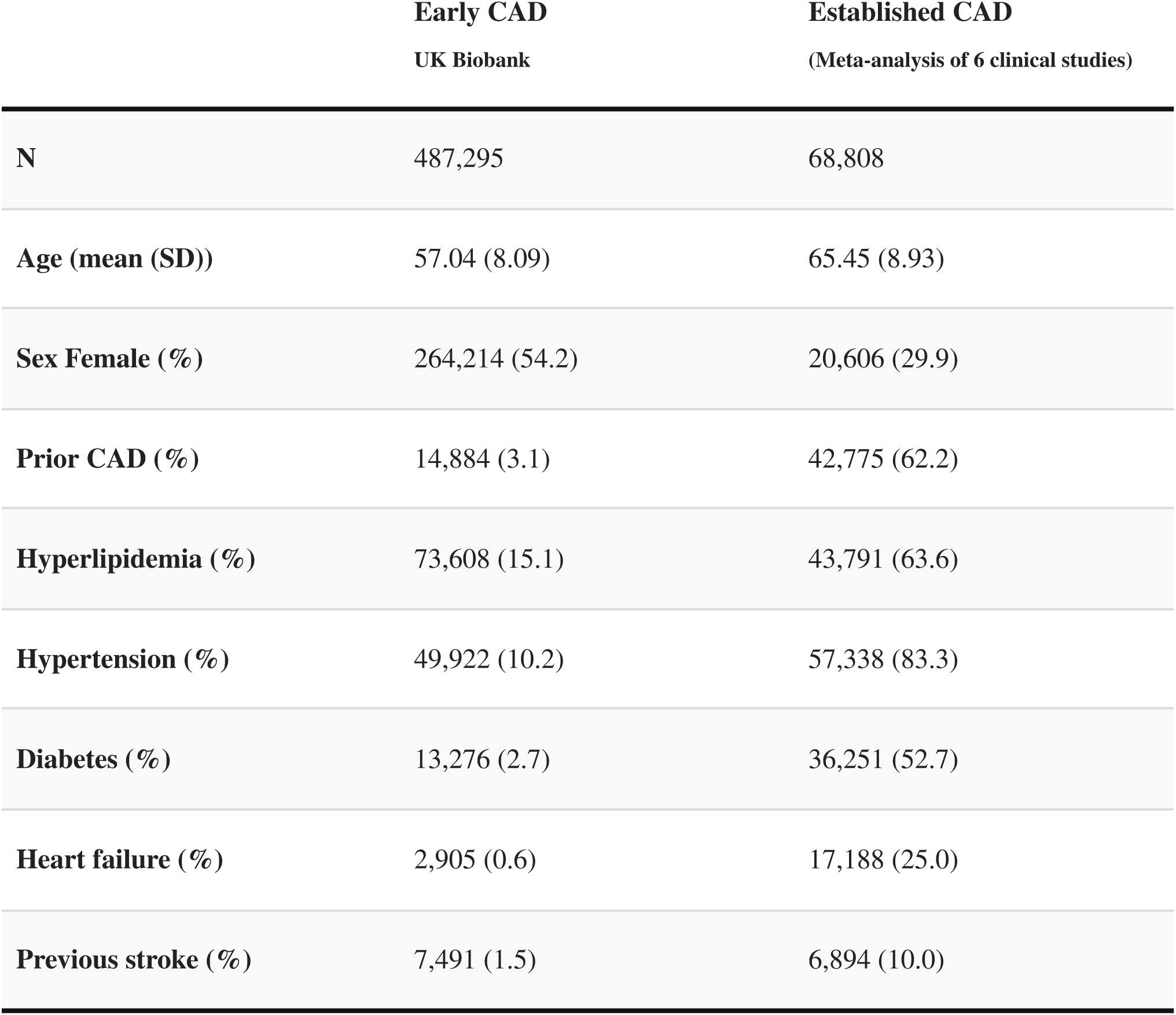
Demographic characteristics of UKBiobank and TIMI trial participants.

|  | Early CAD<br>UK Biobank | Established CAD<br>(Meta-analysis of 6 clinical studies) |
| --- | --- | --- |
| N | 487,295 | 68,808 |
| Age (mean (SD)) | 57.04 (8.09) | 65.45 (8.93) |
| Sex Female (%) | 264,214 (54.2) | 20,606 (29.9) |
| Prior CAD (%) | 14,884 (3.1) | 42,775 (62.2) |
| Hyperlipidemia (%) | 73,608 (15.1) | 43,791 (63.6) |
| Hypertension (%) | 49,922 (10.2) | 57,338 (83.3) |
| Diabetes (%) | 13,276 (2.7) | 36,251 (52.7) |
| Heart failure (%) | 2,905 (0.6) | 17,188 (25.0) |
| Previous stroke (%) | 7,491 (1.5) | 6,894 (10.0) |

We next assessed whether the association in TIMI participants could be explained by clinical risk factors, trial differences, or collider bias. After additional adjustment for hypertension, LDL-C, diabetes, smoking, and treatment, the TIMI estimates remained significant (OR 1.099, 95% CI 1.028-1.175, P=0.0054). Because conditioning on disease status can introduce collider bias (*50*), we applied SlopeHunter (*51*) correction, and the association in TIMI participants was not attenuated (OR 1.206, 95% CI 1.128-1.289, P<0.001). Per trial estimates were at or above 1 in all four trials that did not restrict enrollment to people with prior coronary events (ORs 1.00-1.13, pooled OR 1.06, 95% CI 1.00-1.12), with no detectable between-trial heterogeneity (Cochran’s Q = 2.75, df = 3, *P* = 0.43, I² = 0%, **fig. S17B**).

Finally, we examined subsequent CAD or coronary events (revascularization/MI/CHD death) using Cox regression. Among UKBiobank participants without prior CAD, myocardial infarction or stroke at baseline, V53I was associated with a lower risk of incident CAD over a median follow-up of 11.1 years (HR 0.93, 95% CI 0.89-0.97, P=0.0016, **Fig. 7B, top**). In contrast, among TIMI trial participants, the V53I allele was associated with a higher incidence of new coronary events over a median follow-up of 2.5 years (HR 1.08, 95% CI 1.01-1.15, *P*=0.025, **Fig. 7B**, **bottom**). The difference between UKBiobank and TIMI participants was significant (ratio of HRs 1.16, 95% CI 1.07-1.26, P_interaction_ = 1.7×10^-4^). While differences between human cohorts, such as disease burden and treatments, prevent direct mapping of murine atherosclerosis stages to human disease progression, these contrasting associations are consistent with the context-dependent effects observed in mice.

## DISCUSSION

We recently linked the CCM signaling pathway to five EC transcriptional programs associated with CAD risk using a large CRISPR-interference screen (*2*, *3*, *5*). That study predicted that decreased CCM2 function would be protective against the atherosclerosis underlying CAD. By contrast, strong loss-of-function variants in CCM2 promote CCM disease (*9*, *10*, *14*, *52*, *53*). Here we provide evidence from human genetic studies, knockout ECs, and *Ccm2* heterozygous knockout mice to link CCM2 to stage-specific effects on atherosclerosis (Fig. 8). At baseline, ECs in *Ccm2^+/-^* aortas exhibited an anti-inflammatory transcriptional phenotype. This transcriptional effect is reflected in CCM2KO human ECs, which also show reduced monocyte adhesion and coagulation factor activation. This baseline phenotype likely contributes to reduced early atherosclerosis (12 weeks of high fat diet). However, our scRNA-seq data from the aortas of mice exposed to 12 weeks of high fat diet show a cellular phenotype switch from anti- to pro-inflammatory, particularly in ECs, along with increased circulating TNFα. This pro-inflammatory state likely contributes to a reversal of the *Ccm2^+/-^* effect on atherosclerosis, resulting in increased plaque area at 20 weeks. Human genetic data for the lead *CCM2* variant (rs2107732) shows a similar reversal. It decreases risk of CAD in the general population (UK-BioBank, likely reflecting early inhibition of atherogenesis) but increases risk in patients pre-selected for high risk or established CAD (TIMI trial meta-analysis, consistent with exacerbation of existing atherosclerosis).

**Fig. 8.**
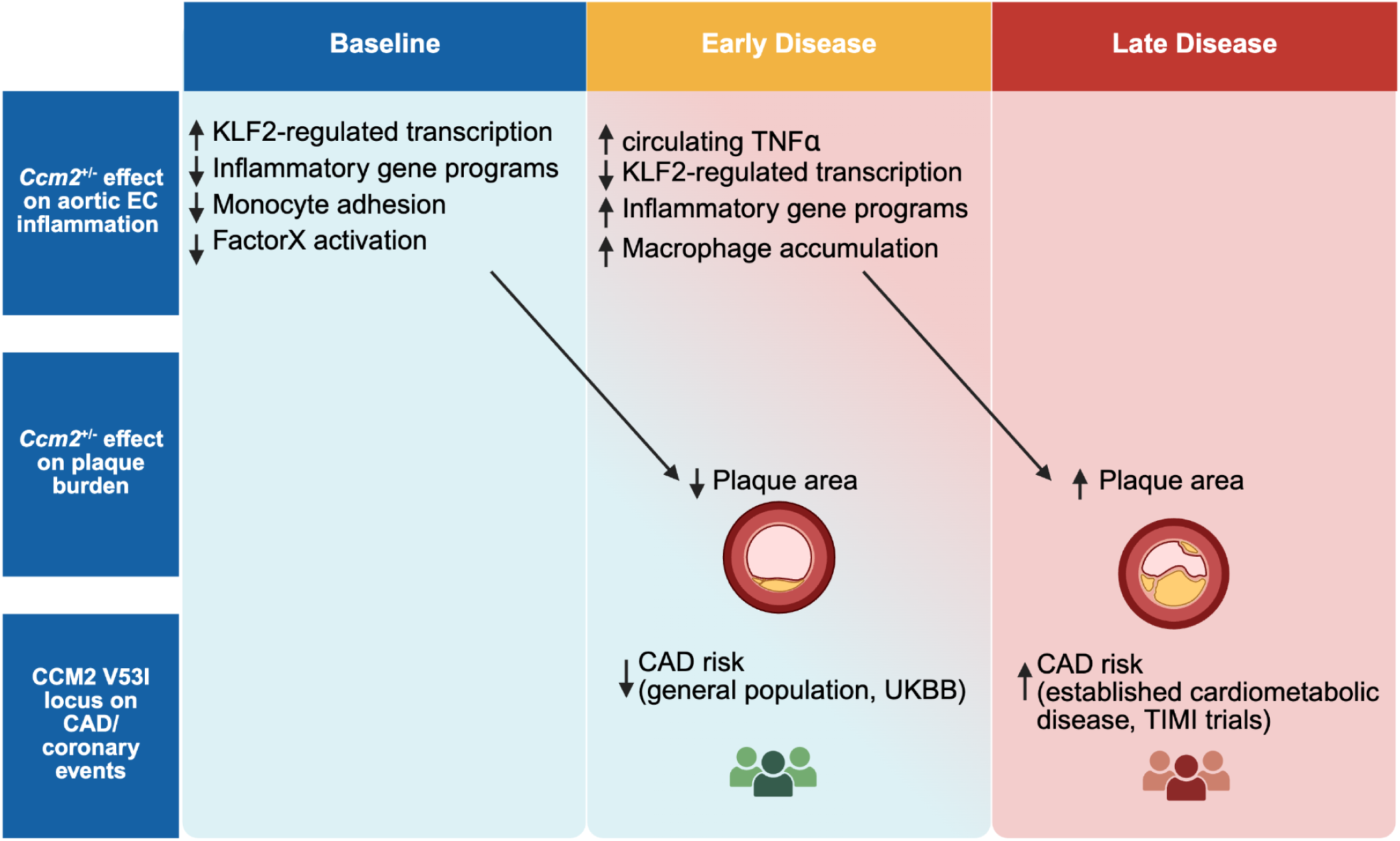
Proposed model of *CCM2* loss-of-function in atherosclerotic vascular disease.

An unexpected finding of this study was that the cellular response to reduced Ccm2 changed with disease progression, across all major aortic cell types, but most strikingly in ECs. At baseline, ECs with reduced *Ccm2* expression exhibited a transcriptional state consistent with athero-protection, with elevated *Klf2* expression and suppression of inflammatory pathways both *in vivo* and *in vitro*. By contrast, after 12 weeks of high fat diet, *Klf2* and its associated protective genes were suppressed (specifically in ECs, out of all major vascular cell types), whereas *NF-κB, STAT,* and IRF-associated inflammatory programs were increased. This switch was accompanied by an increase in circulating TNFα, and we found that TNFα treatment of cultured ECs could eliminate the increased *KLF2* expression caused by CCM2 knockdown, suggesting that high TNFα may promote the switch to pro-inflammatory ECs *in vivo*.

In unstimulated human aortic ECs, conditioned medium (CM) from CCM2KO cells reduced monocyte adhesion, which is consistent with the less inflammatory phenotype observed at baseline in *Ccm2*^+/-^ mouse aortic ECs. In contrast, CM from TNFα stimulated CCM2KO ECs increased monocyte adhesion compared to CM from TNFα stimulated control ECs, consistent with increased cytokine gene expression in mouse aortic ECs at 12 weeks. Interestingly, genes for the adhesion molecules required to tether immune cells to ECs remained low in Ccm2^+/-^ ECs at 12 weeks in vivo, suggesting that the switch to a pro-inflammatory state is not absolute. Correspondingly, we find that CCM2KO ECs showed less monocyte adhesion than controls when treated with conditioned media from control cells stimulated either with or without TNFα. The expression of *Tnf* itself is significantly increased in ECs at 12 weeks, but it is unclear whether this is sufficient to cause the rise of circulating TNFα, or whether that comes from other cells in the aortic wall (T cells also increase *Tnf* expression) or even from other tissues.

While some effects of CCM2 loss-of-function were associated with the classic signaling pathway (of central importance in CCM disease) in which CCM2 inhibits the MEKK3/ERK5 signaling that upregulates KLF2, others were independent of this pathway. In particular, treatment with an ERK5-degrading PROTAC reversed the reduced binding of monocytes to CCM2KO ECs but did not block reduced Factor X activation in CCM2KO ECs. This latter effect may be because of the ERK5-independent downregulation of Coagulation Factor III (F3), an activator of this process, in CCM2KO ECs. The CCM complex also signals through RhoA-ROCK1/2 and PI3 kinase (*12*), and future studies will be needed to discern if these, or some novel signaling process, mediates the effect of CCM2 loss on Factor X activation. Nonetheless, these observations serve as a reminder that individual aspects of complex signaling mechanisms which are relevant to one disease may not be the only ones that are important for other disease processes.

One limitation of this study is that the whole body *Ccm2*^+/-^ heterozygous mouse model cannot positively identify which cell types are necessary or sufficient for the observed effects on atherosclerosis. We used *Ccm2*^+/-^ gene-trap mice that have previously served to identify roles for Ccm2 in cytoskeletal architecture, barrier function, and angiogenesis in brain ECs (*14*, *53*). We chose this model because 1) partial loss of function is generally expected to be a better model, relative to complete knockout, of the effects of common variants on disease, and 2) whole body knockout better mirrors the general effects of germline variants in humans, and also allows us to consider the effects of reduced Ccm2 function on all major arterial cell types. The results of these studies suggest that ECs might be sufficient to drive both the early decrease and late increase of atherosclerosis, and future studies using temporally controlled, cell type-specific Ccm2 deletion will be needed to test this hypothesis - as well as to examine whether the transcriptional effects of *Ccm2*^+/-^ that we see in other cell types also modulate the atherosclerosis phenotype. Another limitation of this study is that, while it links the V53I variant in CCM2 to context-specific effects on CAD risk, and links reduced Ccm2 function to context-specific effects on atherosclerosis in mice, it does not identify the specific mechanism by which the V53I variant affects disease. Future studies to identify this mechanistic link will need to overcome two challenges that are pervasive in the field: 1) that, to not be strongly selected against, common disease-associated variants often have small effect sizes that may take decades to manifest *in vivo*, and 2) the effects of these variants are likely to appear only in complex disease contexts that may be difficult to reproduce *in vitro*.

Our data highlight a growing debate about the importance of GWAS findings in disease initiation versus progression. A recent study which analyzed data from seven longitudinal biobanks for 9 common diseases found limited overlap between the variants that affect disease risk and those associated with survival (*54*). For CAD in particular, the variants with a significant effect on disease susceptibility had an inverse correlation with disease-specific mortality. In other studies, the prototypical CAD 9p21 GWAS locus tracks with the presence and early growth of atherosclerotic burden but shows a weaker or more heterogeneous relationship with plaque evolution and rupture in longitudinal cohorts (*55*, *56*). These paradoxical findings could be attributed to collider bias, since individuals with disease, despite low genetic risk, may have high exposure to other unmeasured risk factors. Our data showing that *Ccm2* heterozygosity switches from reducing early atherosclerosis to increasing late stage atherosclerosis, however, provides support for a possible true biological reason for the incongruity between the genetics of CAD initiation and progression.

In summary, our studies of mouse atherosclerosis, human EC function, and human genetics identified *CCM2* as a context-specific regulator of atherosclerosis: with *Ccm2* heterozygosity decreasing early atherosclerosis but increasing late stage atherosclerosis, and also with the lead human *CCM2* variant having opposing effects on CAD in low versus high risk populations. The observation that early atherosclerosis was reduced in *Ccm2^+/-^* mice, with ECs likely playing a key role in this effect, partially validated the hypothesis arising from our Perturb-seq study, and supported the value of our variant-to-gene-to-program approach. The switch to increased late stage atherosclerosis, however, indicates that a screen under a single condition *in vitro*, despite identifying valid disease-associated genes, may miss important nuances of those genes’ roles in disease initiation and progression. Our results indicate that therapeutically targeting CCM signaling has the potential to help in the prevention or treatment of CAD, but only when therapeutics are matched to the correct stages of disease. They also emphasize caution for therapeutics in development for cerebral cavernous malformations, given that our findings suggest that compounds increasing CCM functionality in brain microvascular ECs may have unintended consequences in other vascular beds. More generally, our results emphasize the importance of integrating human genetic data from cohorts at different stages of disease for nominating potential new drug targets, and highlight that all GWAS-supported drug targets should be carefully studied for protective effects at multiple stages of disease.

## MATERIALS AND METHODS

### Study design

#### Mouse studies

We performed atherosclerosis studies in *Ccm2*^+/-^ mice and *Ccm2*^+/+^ littermates. Cohort sizes were calculated by power analysis (*57*) to detect a 10% difference in aortic root lesion area with 80% power and were consistent with prior studies also using the AAV-PCSK9 models in the same facility (*58*, *59*). Mice of both sexes were enrolled as available from littermate breeding, without selection by sex. Mice with plasma cholesterol level comparable to uninjected mice, indicating failed AAV-PCSK9 injection, were excluded (a total of one mouse in this study). Histological staining and analyses were conducted by researchers blinded to genotype.

#### Human genetics

We used statistical fine-mapping and colocalization of the *CCM2* locus to prioritize the most likely causal variant. Because CAD GWAS was a meta-analysis and LD was estimated from an external reference panel, discrepancies between the association statistics and the reference LD can produce miscalibrated results (*60*). Thus, we prioritized CARMA (*40*), a Bayesian fine-mapping method that explicitly accommodates inconsistencies between summary statistics and the LD reference. We then tested the association of the lead variant (rs2107732) in the *CCM2* locus with CAD in two independent populations: UKBiobank and TIMI clinical trials. Cross-sectional association was done using logistic regression and the prospective analysis of subsequent CAD was performed using Cox regression. Sensitivity was tested through covariate-adjusted models, trial-level analysis, and collider bias correction.

### Mice

The global *Ccm2*^+/-^ mouse model in a C57BL/6J background that was used in this study was a generous gift from Dr. Douglas Marchuk at Duke University. In this mouse, the *Ccm2* heterozygous knockout was achieved via gene-trap insertion in Exon 6 (*14*). Animal maintenance and experimental procedures were approved by the Institutional Animal Care and Use Committee at the Brigham and Women’s Hospital, following the recommendations on animal atherosclerosis from the American Heart Association (*61*). *Ccm2*^+/-^ mice and the wildtype littermate *Ccm2*^+/+^ mice, both male and female, were used in this study. At 10 weeks of age, mice were injected with AAV2/8-PCSK9-D377Y (5x 10^11^ viral genomes, generated by the Viral Core at Boston Children’s Hospital) through tail vein injections. The mice were then subjected to a high-fat, high-cholesterol diet (D12108Ci, Research Diets, Inc.) for 12 weeks or 20 weeks before sacrifice. After euthanasia, blood samples were collected using cardiac puncture and plasma were collected through centrifugation (4°C, 10,000 g, 10 min). Total cholesterol was measured using a colorimetric total cholesterol kit (999-02601, Fujifilm). Plasma cytokine levels were quantified using a Luminex xMAP bead-based multiplex immunoassay by EVE technologies (the Mouse Focused 10-plex, Calgary, AB, Canada).

### Histological analysis of atherosclerotic lesions

After dissection, aortic roots were embedded in Optimal Cutting Temperature (OCT) compound. Cryosections were collected at six different locations along the aortic root (every 42 μm starting from the base of the aortic root). Atherosclerotic lesions were revealed by Oil Red O staining accompanied by hematoxylin counter staining as in previous publications (*62*, *63*). Similarly, the aortic arch, along with the brachiocephalic artery (BCA), left common carotid artery, and left subclavian artery, was embedded in OCT. Three coronal sections, where both the aortic arch and the BCA were visible, were subjected to Oil Red O staining to detect atherosclerotic lesions. Histological images were acquired using Olympus slide scanners (VS120, VS200) under 20x magnification. Fiji/ImageJ was used for image analysis. At the aortic root, areas of atherosclerotic lesions were measured. Necrotic core area in the same sections was quantified as regions lacking intact cells and may contain cellular debris or cholesterol crystals (*61*). For CD68 immunostaining, OCT-embedded frozen aortic root sections were fixed in methanol for 20 min at −20°C, rehydrated in water for 5 min, washed (K800621, Dako), and permeabilized (39487, CST) for 5 min at room temperature. Sections were then blocked (K800721, Dako) for 1 h at room temperature and incubated overnight at 4°C with anti-CD68 antibody (1:100; 977783, CST). After washing, sections were incubated with Alexa Fluor 555-conjugated goat anti-rabbit secondary antibody (1:500) for 1 h, counterstained with Hoechst 33342 (1:1,000) for 5 min, and mounted. Collagen deposition was quantitated using Masson’s Trichrome stain following manufacturer’s instruction (ab150686, Abcam). All sections were imaged using an Olympus VS200 automated slide scanner at the Harvard MicRoN Core. Additionally, the descending aortas were dissected and fixed in 10% neutral buffered formalin overnight. Aortic arches and descending aortas from mice after 20 weeks of atherogenic diet were harvested and fixed in 10% formaldehyde (HT501128, Sigma) overnight and subjected to *en face* Oil Red O Staining. In brief, aortas were incubated with propylene glycol for 15 min (822324, Sigma) and then stained with Oil Red O solution for 30min (26079-15, Electron Microscopy Sciences) under room temperature. The samples were washed with propylene glycol and then water to remove the excess dye, and differentiated in water overnight at 4 ℃. All samples were processed by researchers who were blinded to the experimental design.

### Mouse aorta single cell RNA-seq

Mouse aortas (ascending, arch, and descending) were dissected and incubated at 37°C in the enzymatic digestion buffer for 40min. Enzymatic digestion buffer recipe: Collagenase I (450 U/ml, C0130, Sigma-Aldrich), Collagenase XI (125 U/ml, C7657, Sigma-Aldrich), DNAse I (60 U/ml, 11284932001, Sigma-Aldrich), Hyaluronidase (60 U/ml, H3506, Sigma-Aldrich), HEPEs (20 mM, 15630080, Gibco) in 10%FBS-PBS. Next, the tissue suspension was passed through a 70 μM cell strainer and red blood cells were removed using ACK lysis buffer (A1049201, Gibco). For each genotype-by-timepoint group, aortas from n = 3-4 (n=4 for both genotypes in Week 0, n=3 for both genotypes in Week 12) mice were harvested. Each aorta was enzymatically dissociated separately and an equal number of viable cells from each mouse in the same genotype x timepoint group was then combined to generate the cell suspension used for sorting. The cell suspensions were subjected to CD31 (558738, BD Pharmingen) and 7-AAD viability (00-6993-50, eBioscience) staining. Live CD31+ and CD31-cells were sorted into separate tubes using a FACSAria cell sorter (BD Biosciences). Both fractions were retained for sequencing, with approximately equal number (10k) of cells from each fraction loaded for 10x reactions. This sampling strategy was used to ensure sufficient EC recovery while retaining non-EC populations. Because the two sorted fractions were sampled independently, cell type ratios in the resulting dataset were not interpreted as estimates of absolute cellular abundance in the aorta. Library preparation was performed following the manufacturer’s instruction (Chromium Next GEM Single Cell 3’ GEM, Library & Gel Bead Kit v3.1, 10x Genomics). Libraries were sequenced on NextSeq 500 aiming for 20,000 reads per cell. Count matrices were obtained using the CellRanger (Cloud Analysis, 10x Genomics) pipeline and the reads were aligned to the mouse reference genome Mm10-2020-A.

### Single cell RNA-seq analysis

Single cell RNA-seq data were analyzed using R (version 4.3.1) and can be found at (GSE315884). The analysis pipeline has been deposited on GitHub (https://github.com/HouseFang-Broad/CCM2hetKOmice_aorta_scRNAseq). Ambient RNA was removed using the CellBender v0.3.2 with default parameters, except that a learning rate of 5×10^-5^ was used. Quality control was then performed using scanpy v1.10.3, and droplets meeting the following criteria were preserved in the count matrices: log_10_genesperUMI>0.8, cells with min_counts=500, cells with min_genes=250, genes with min_cells=10, and cells with percent mitochondria gene expression < 20%. Potential doublets were determined using the scanpy.pp.scrublet function (using default parameters with an expected_doublet_rate=0.1). Cells with doublet_score>0.4 were deemed as potential doublets. Harmony (*64*) was used for data integration. Marker genes of each cluster were determined using the FindAllMarkers function from the R package Seurat V4 (*65*). Cell types were annotated based on markers identified from Panglao (*66*) and previous publications (*20*, *67*). The pooling of bioreplicates of the same group limits the statistical analyses possible for cell type fractions. We therefore used scProportionTest (*36*), a permutation-based proportion test with bootstrap confidence intervals to describe changes in cell fractions. Differential expression was performed on the RNA assay using the Wilcoxon rank-sum test (Seurat). Tests were run separately within each cell type at each timepoint, comparing *Ccm2*^+/-^ versus *Ccm2*^+/+^. Differentially expressed genes were deemed as those with a Benjamini-Hochberg adjusted *P* value lower than 0.05 and minimum absolute log_2_FC of 0.2. Pathway enrichment was performed with clusterProfiler v4.10.1(*68*) against three gene-set collections retrieved with msigdbr v25.1.0 (*69*, *70*) for Mus musculus: Gene Ontology Biological Process (C5, GOBP), KEGG (C2, CP:KEGG_LEGACY) and Reactome (C2, CP:REACTOME). Each contrast was one cell type at one timepoint (*Ccm2*^+/-^ versus *Ccm2*^+/+^). For each contrast, genes with a nominal Wilcoxon *P* < 0.05 were retained and ranked by sign(average log_2_FC) × (-log_10_(P)). Gene set enrichment analysis was run on this ranked list with clusterProfiler::GSEA (fgsea, v1.28.0). Gene sets containing fewer than 15 or more than 1,000 genes were not tested. Enrichment *P* values were corrected by Benjamini-Hochberg within each cell type and database, and pathways with Padj ≤ 0.05 were considered significant. To infer transcription factor activities from DE gene sets, we utilized decoupleR with DoRothEA regulons (*31*).

Genes of interest representing key biological functions of a cell type’s contribution to atherosclerosis are selected based on published literature. ECs: (*20*, *22*, *71*) (*72–80*). Macrophages: (*67*, *81–87*). VSMCs: (*67*, *88–90*). Fibroblasts: (*91–94*).

### Cells

Telomerase-immortalized human aortic ECs (teloHAECs) were purchased from ATCC (CRL-4052) and maintained in a low serum medium for human ECs (LL-0003, Lifeline). CCM2 knockout in teloHAECs was achieved by delivering the ribonucleoprotein complex containing SpCas9 nuclease and three sgRNAs targeting human CCM2 (Gene knockout kit, Synthego) via nucleofection (program CA210, AAF-1003B, Lonza) as described before (*3*), using non-targeting guides (NTC1) for control lines. Clonal knockout and control lines were isolated by serial dilution. CCM2 knockout was confirmed by Sanger sequencing and next generation sequencing. PROTAC INY-06-061 and the control PROTAC INY-06-089 were generous gifts from Dr. Stephen MacLeod Hinshaw. siRNA-mediated CCM2 knockdown was performed using reverse transfection (13778075, Invitrogen). CRISPRi-mediated *CCM2* knockdown and Perturb-seq analysis followed our previous publication (*3*).

### Quantitative PCR (qPCR)

RNA was isolated using the RNeasy kit (74104, Qiagen) and genomic DNA was removed by incubation with DNase (12185010, Invitrogen). 400 ng RNA was used for reverse transcription following the manufacturer’s instruction (4374967, Applied Biosystems). qPCR was conducted on QuantStudio5 (A34322, Applied Biosystems). The following Taqman probes were used to measure mRNA expression in either mouse aortas (Ccm2, Mm01183004_g1) or cultured human aortic ECs (*CCM2* Hs01123851_g1; *KLF2* Hs07291763_gH, *KLF4* Hs00358836_m1, *F3* Hs01076029_m1, *TFPI* Hs00409206_m1). GAPDH for mouse (Gapdh, Mm99999915_g1) or human (GAPDH, Hs02786624_g1) were used as housekeeping genes. Relative expression was assessed by calculating the 2^-ddCT^.

### Monocyte adhesion assay

TeloHAECs were treated with recombinant human TNFα at 10 ng/ml (300-01A, Peprotech) overnight prior to the day of assay. Human monocyte cell line THP-1 (TIB-202, ATCC) was labeled with calcein AM (C3099, Invitrogen) for 30 min, then washed with PBS twice before incubation with teloHAECs. After 45 mins of incubation, unbound labeled THP-1s were washed away and the adhered cell suspension containing both teloHAECs and THP-1s was prepared by trypsin digestion. The total number of THP-1 adhered to teloHAEC was quantified by flow cytometry (CytoFLEX, Beckman Coulter). Efficiency of teloHAEC monocyte recruitment was assessed by the number of THP-1s adhered to every 1,000 teloHAECs to avoid potential artifacts introduced by plating density and proliferation rate. For conditioned media assays, CCM2KO or WT teloHAECs were seeded at 2×10^5^ cells per well in 6-well plates and cultured for two days, followed by treatment with TNFα (10 ng/mL) for 20 h or no treatment. Conditioned media were collected and transferred to recipient teloHAECs after removal of their original culture media. Monocyte adhesion assays were performed 4 h after conditioned media transfer.

### Bulk RNA-seq

RNA from cultured aortic ECs was isolated as described in the qPCR section. After genomic DNA removal, 400ng of RNA was used for library preparation. mRNA was isolated by polyA selection (E7490S, New England Biolabs) and the RNAseq libraries prepared as per manufacturer’s instructions (E7770S, New England Biolabs). Libraries were sequenced on NextSeq500 (75 cycles, pair-ended reads) and demultiplexed using the Walk-Up Sequencing service at the Broad Institute. Sequencing results were mapped to the human hg19 genome build using STAR. Downstream analysis was conducted using edgeR (*95*) according to the user’s manual. Data can be found here: teloHAEC with CRISPRi-mediated CCM2 knockdown: GSE232400 (GSM7329970, GSM7329977, GSM7329967, GSM7329974, GSM7329962, GSM7329959, GSM7329968, GSM7329965), teloHAEC with CCM2 knockout: GSE316335, GSE262246 (GSM8161046, GSM8161042)).

### Factor X activation assay

TeloHAECs with CCM2 knockout or non-targeting controls were used for a Factor X activation assay as previously described (*96*). Briefly, 2×10^5^ cells were seeded in each well on a 96-well plate overnight. The cells were then stimulated with 10 ng/ml TNFα (R&D Systems) for 3.5 hours at 37°C. Next, the cells were incubated with 125nM Factor X (Prolytix), 0.6nM Factor VIIa (Prolytix), and 150μM Biophen-CS11 chromogenic substrate (Aniara Diagnostica). To quantify Factor X activation, absorbance at 405nm was continuously measured for 3 hours on an xMark Spectrophotometer (Bio-Rad). Maximum reaction velocity (Factor Xa nM) was calculated based on the standard curve established by purified Factor Xa (Prolytix).

### Bayesian fine mapping

For fine mapping the CCM2 locus for causal variants, we followed the established pipeline in previous publications (*40–43*, *97*). GWAS summary statistics were computed from the most recent coronary artery disease GWAS study in over a million individuals (*2*) (GCST90132314). We defined the fine-mapping regions based on a 3 Mb window around each lead variant and merged regions if they overlapped (*97*). The CCM2 region is chr7.43577978-46577978 (hg19). Fine mapping was performed with reference linkage disequilibrium (LD) from the UK Biobank. LD was computed using LDstore v2.0 (*98*) with unrelated “white British” samples from the UK Biobank, QC’ed in the Neale lab GWAS (URL: https://www.nealelab.is/uk-biobank), then further filtered to samples with available CAD phenotype in the UK Biobank (N=360,710). The Locus Zoom plot was generated based on the most recent CAD GWAS meta-analyses (*2*) using LocusZoom(*99*). PheWAS data were obtained from previous systemic PheWAS studies (*100*).

### Colocalization analysis

Colocalization analysis was performed using the Coloc v5.2.2 R package. For input, we used subsetted CAD GWAS summary statistics and human peripheral blood cis-eQTL data, focusing on a 1MB region (±500k bp) centered on the first base pair of the CCM2 coding region. CAD GWAS summary statistics were obtained from the same study mentioned in the Bayesian fine-mapping section. Human peripheral blood cis-eQTL data for CCM2 was obtained from the eQTLGen phase I database. Phase 3 1000 Genomes Project minor allele frequencies paired with the eQTL data were downloaded from the ensembl.org Allele Frequency Calculator reference. To account for variation in SNP meta-analysis sample sizes, the standard deviation of the quantitative trait (sdY) was imputed as per coloc’s standard procedure for sdY estimation. A PP.H4 > 0.8 would be considered as strong evidence for a variant to have colocalization of GWAS risk and CCM2 eQTL.

### Genetic analysis in UK-Biobank and cardiovascular clinical trials

To determine the association between rs2107732 and coronary artery disease (CAD), we used multivariable logistic regression adjusted for age, sex, and population structure (principal components 1–5), and report odds ratios (ORs) with 95% confidence intervals (CIs). In UK Biobank, genotype was modeled by a dominant model comparing A-allele carriers (AA/AG) with GG as the reference. Additionally, the UK Biobank genotype was modeled categorically as AA, AG, or GG, with GG as the reference. In the six clinical-trial cohort, subjects from DECLARE-TIMI 58, ENGAGE AF-TIMI 48, FOURIER, SAVOR-TIMI 53, SOLID-TIMI 52, and PEGASUS-TIMI 54 were pooled, and rs2107732 was analyzed under a dominant model. The clinical-trial outcome combined both incident and prevalent CAD, including any history of CAD, myocardial infarction, or percutaneous coronary intervention, and therefore predominantly reflecting disease present before trial enrollment. For further adjustment for cardiovascular risks, we used a sensitivity model additionally adjusted for hypertension, LDL cholesterol, diabetes, and smoking. Analyses incorporating LDL cholesterol were restricted to participants with available covariate data. To test whether the association of V53I differed between UKBiobank and TIMI participants (the interaction test), we compared the corresponding log(OR or HR) estimates from the two cohorts using a two-sided Wald test, with the standard error of the difference calculated as the square root of the summed variances (*101*).

Prospective associations were evaluated using Cox regression under the dominant genetic model and are reported as hazard ratios (HRs) with 95% CIs. In UK Biobank, participants with CAD, myocardial infarction, or stroke at baseline were excluded, and incident CAD was assessed over a median follow-up of 11.1 years. In the six clinical trials, 61,047 participants with follow-up data were evaluated for incident coronary revascularization, myocardial infarction, or coronary heart disease death after enrollment over a median follow-up of 2.5 years. Analyses were stratified by baseline cardiovascular status as high risk with/without clinically manifest atherosclerotic cardiovascular disease (ASCVD).

For trial-specific sensitivity analyses, dominant-model ORs were estimated separately in the four trials that had both CAD cases and non-cases (DECLARE-TIMI 58 (*44*), ENGAGE AF-TIMI 48 (*45*), FOURIER (*46*), and SAVOR-TIMI 53 (*47*)) and combined by inverse-variance meta-analysis on the log-OR scale. Between-trial heterogeneity was assessed using Cochran’s Q and I². SOLID-TIMI 52 (*48*) and PEGASUS-TIMI 54 (*49*) did not contribute to this analysis because the CAD phenotype provided no within-trial case–control comparison in these cohorts. Differences between independent prospective estimates were tested on the logHR scale using the difference in coefficients and the sum of their variances. Analyses were performed in R version 4.4.1.

### Statistical analysis

Bioinformatic analyses of Bayesian fine-mapping, bulk RNAseq, and scRNAseq have been described in the prior sections. For animal studies and cellular assays, all data were presented as mean±SEM and statistical analyses were performed in GraphPad Prism 10. Comparisons between two independent groups were analyzed using the t test. Comparisons involving multiple groups or two independent variables (e.g. genotype and sex) were analyzed with one-way ANOVA with Tukey’s post hoc test or two-way ANOVA with Šídák’s multiple comparisons, indicated in the figure legends. A *P* value<0.05 was deemed statistically significant.

## Data Availability

All data produced in the present study are available upon reasonable request to the authors.

## Acknowledgments

We’d like to thank Dr. Douglas Marchuk for generously sharing the *Ccm2*^+/-^ mice, and Dr. Nathanael Gray and Dr. Stephen Hinshaw for providing ERK5 PROTAC INY-06-061.

## Author contributions

Conceptualization: SF, RG

Experimental design and execution: SF, AV, NP, MC, AG, CZ, RM

Data analysis: SF, GS, CZ, SMZ

Clinical study data and analysis: JJ, FK, CR, MS, NM

Statistical fine-mapping: RC

AlphaFold modeling: MT

Writing – original draft: SF

Writing – review & editing: SF, RG, GS, JE, AS, PL

## Data and materials availability

The sequencing datasets generated in this study are being prepared for deposition in the NCBI Gene Expression Omnibus (GEO). Data supporting the findings presented in this preprint are available from the corresponding author upon reasonable request, pending public repository release.

## Supplementary Materials

**Table S1.** GSEA results at Week 0. Table S2. GSEA results at Week 12.

---

**Fig. S1.**
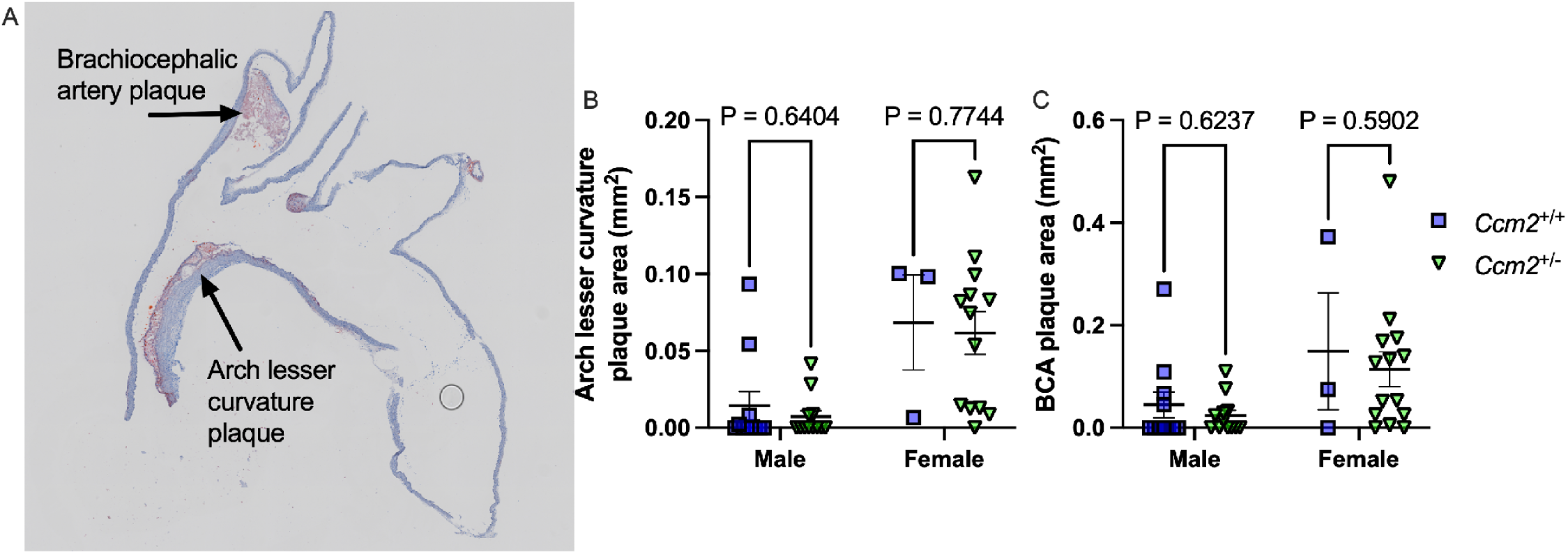
Average plaque area in the aortic arch lesser curvature and brachiocephalic artery. (**A**) Representative image of aortic arch lesser curvature and brachiocephalic (BCA) artery plaques revealed by Oil Red O staining. (**B**) Quantification of aortic arch lesser curvature plaque area (mm²) in male and female *Ccm2*^+/+^ and *Ccm2*^+/-^ mice (P_sex_ = 0.0004, P_genotype_ = 0.6207). (**C**) Quantification of BCA plaque area (mm²) in male and female *Ccm2*^+/+^ and *Ccm2*^+/-^ mice (P_sex_ = 0.0108, P_genotype_ = 0.1812). Statistical tests: Two-way ANOVA with Šídák’s multiple comparisons test to assess P_sex_ and P_genotype_.

**Fig. S2.**
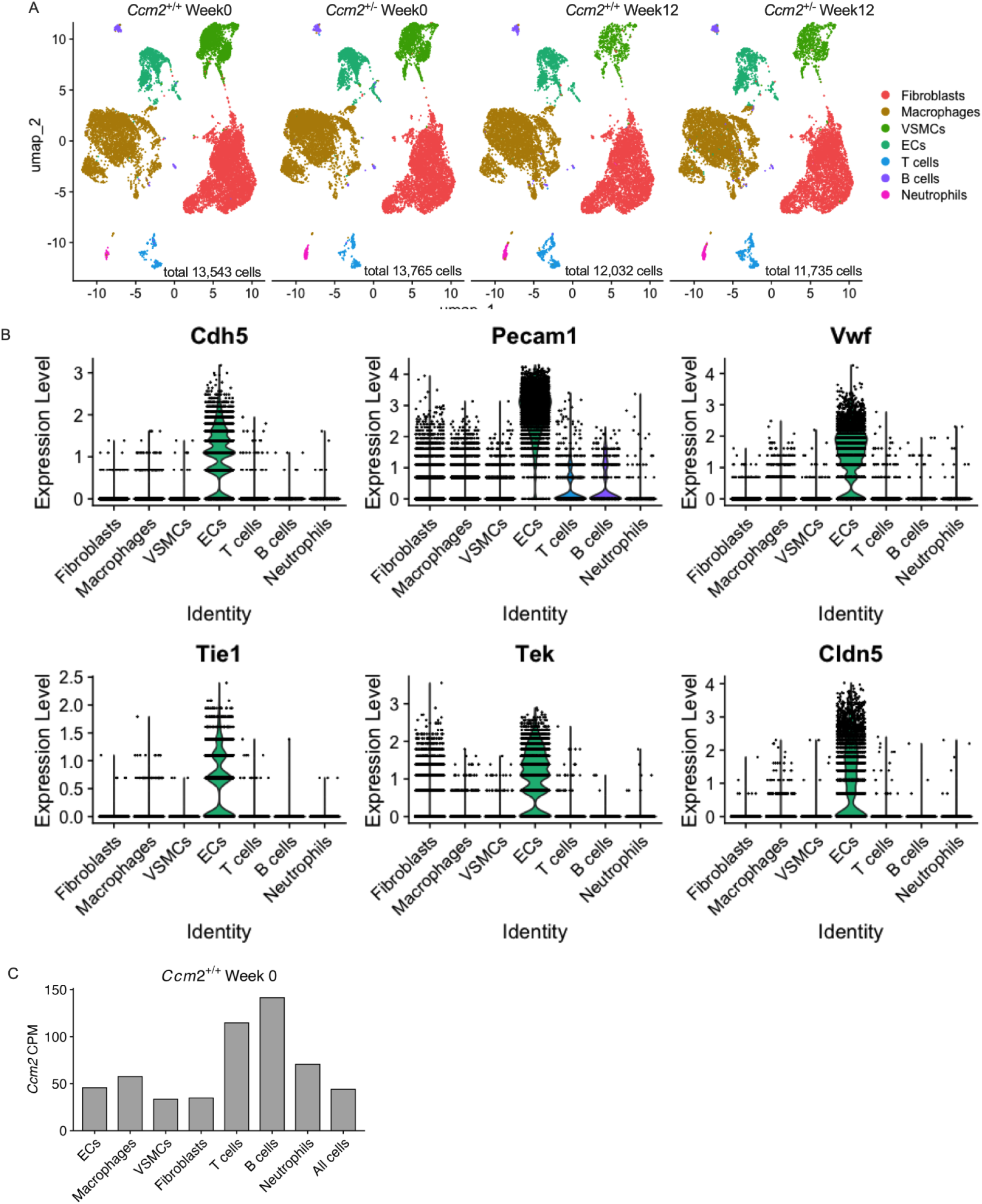
Cell counts and annotations of mouse aortic single cell RNA-seq. (**A**) UMAP showing total cell counts for each combination of genotype and timepoint (*Ccm2*^+/+^ and *Ccm2*^+/-^ mice at Week 0 and Week 12). (**B**) Expression levels of canonical endothelial markers (*Cdh5*, *Pecam1*, *Vwf*, *Tie1*, *Tek*, and *Cldn5*) across major aortic cell types. Each dot represents a cell. (**C**) Ccm2 expression (as counts per million/CPM) in each aortic cell type in *Ccm2*^+/+^ mouse aortas.

**Fig. S3.**
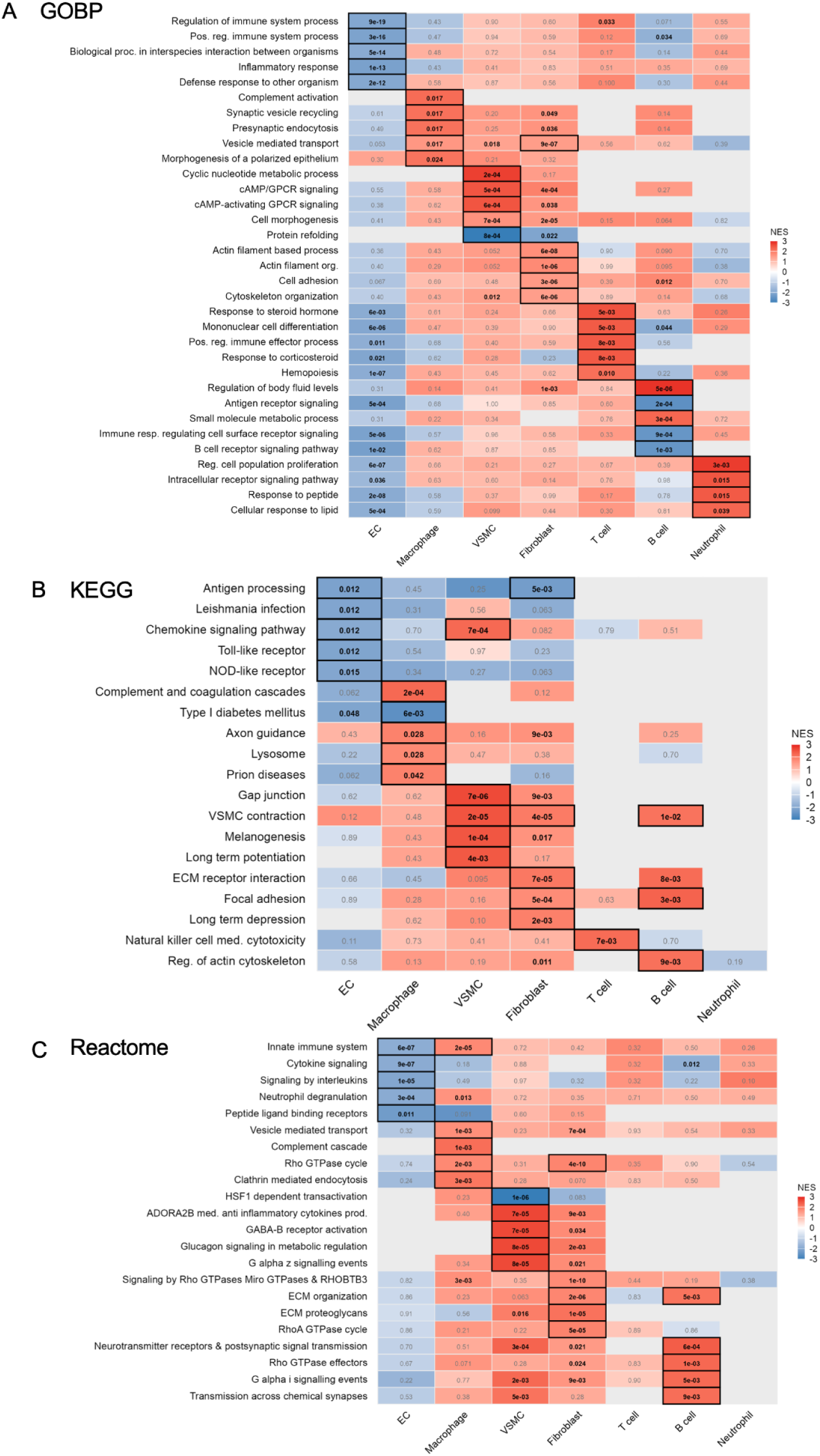
GSEA analysis with GOBP/KEGG/Reactome terms at Week 0. (**A**) Top 5 enriched pathways using GOBP terms, ranked strictly by -log_10_FDR in each aortic cell type *Ccm2*^+/-^ - *Ccm2*^+/+^. (**B**) As in (A), using KEGG terms. (**C**) As in (A), but using Reactome terms. NES, normalized enrichment score. Numbers in boxes, enrichment FDR.

**Fig. S4.**
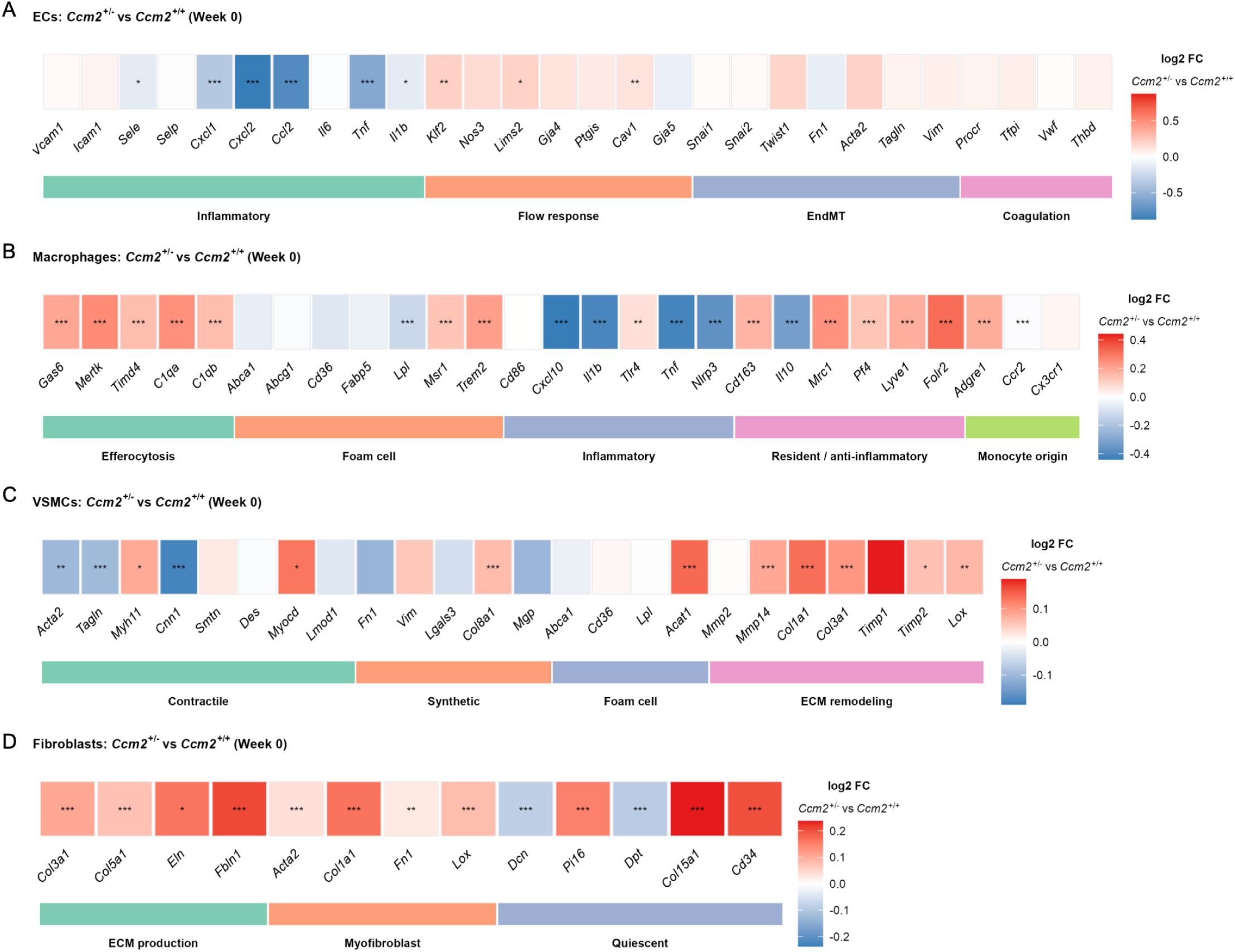
Heatmaps of genes important for atherosclerosis-relevant cellular phenotypes in the 4 most abundant aortic cell types at Week 0. Log_2_FC (*Ccm2^+/-^* versus *Ccm2*^+/+^) of these genes in (A) ECs, (B) Macrophages, (C) VSMCs, and (D) Fibroblasts. *FDR < 0.05, **FDR < 0.01, ***FDR< 0.001.

**Fig. S5.**
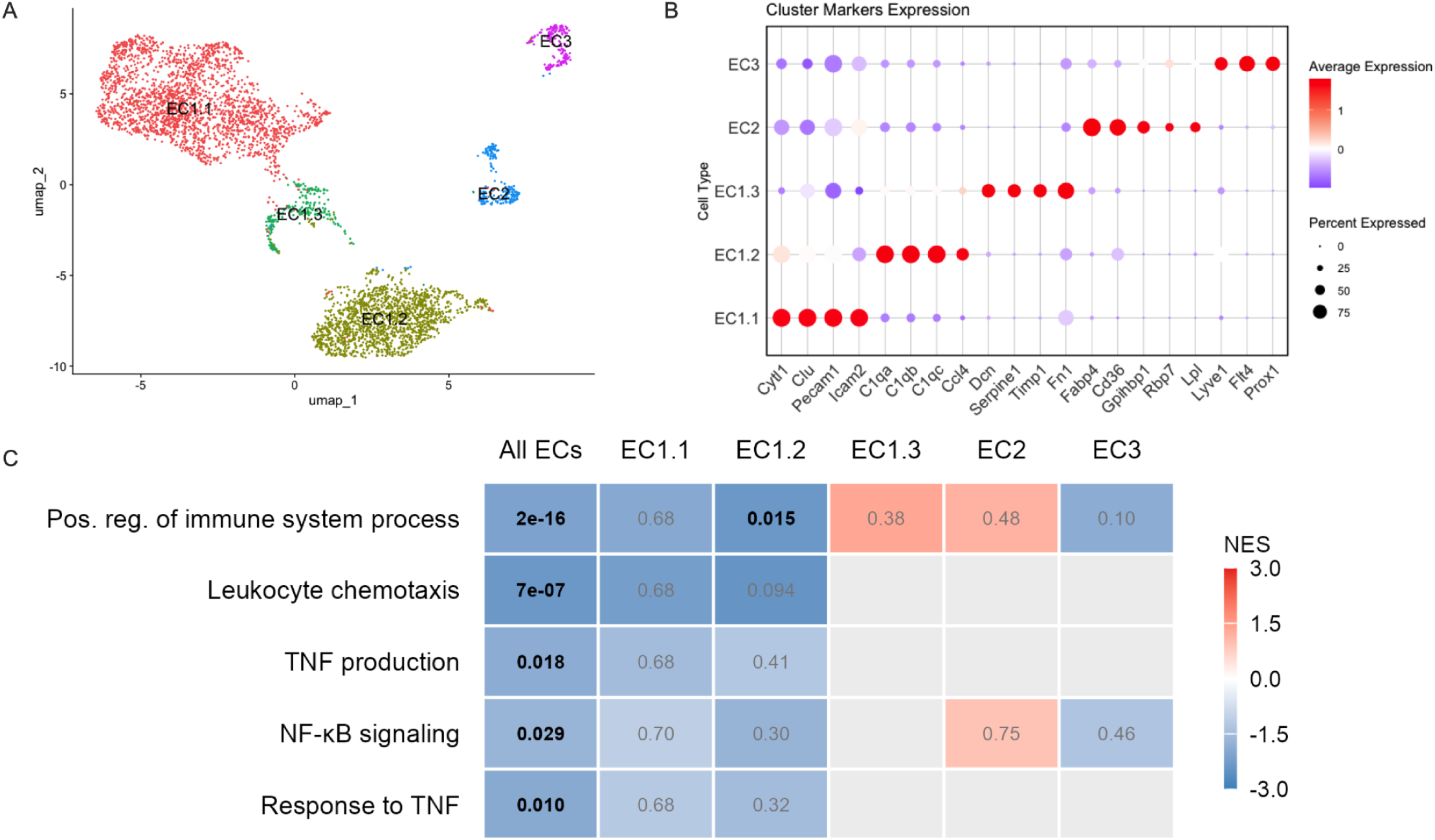
Endothelial cell subcluster analysis at Week 0. (**A**) UMAP of EC subclusters. (**B**) Marker genes for each EC subcluster. (**C**) GSEA normalized enrichment scores (NES) for GOBP pathways in all ECs and individual EC subclusters. Numbers in boxes: FDR.

**Fig. S6.**
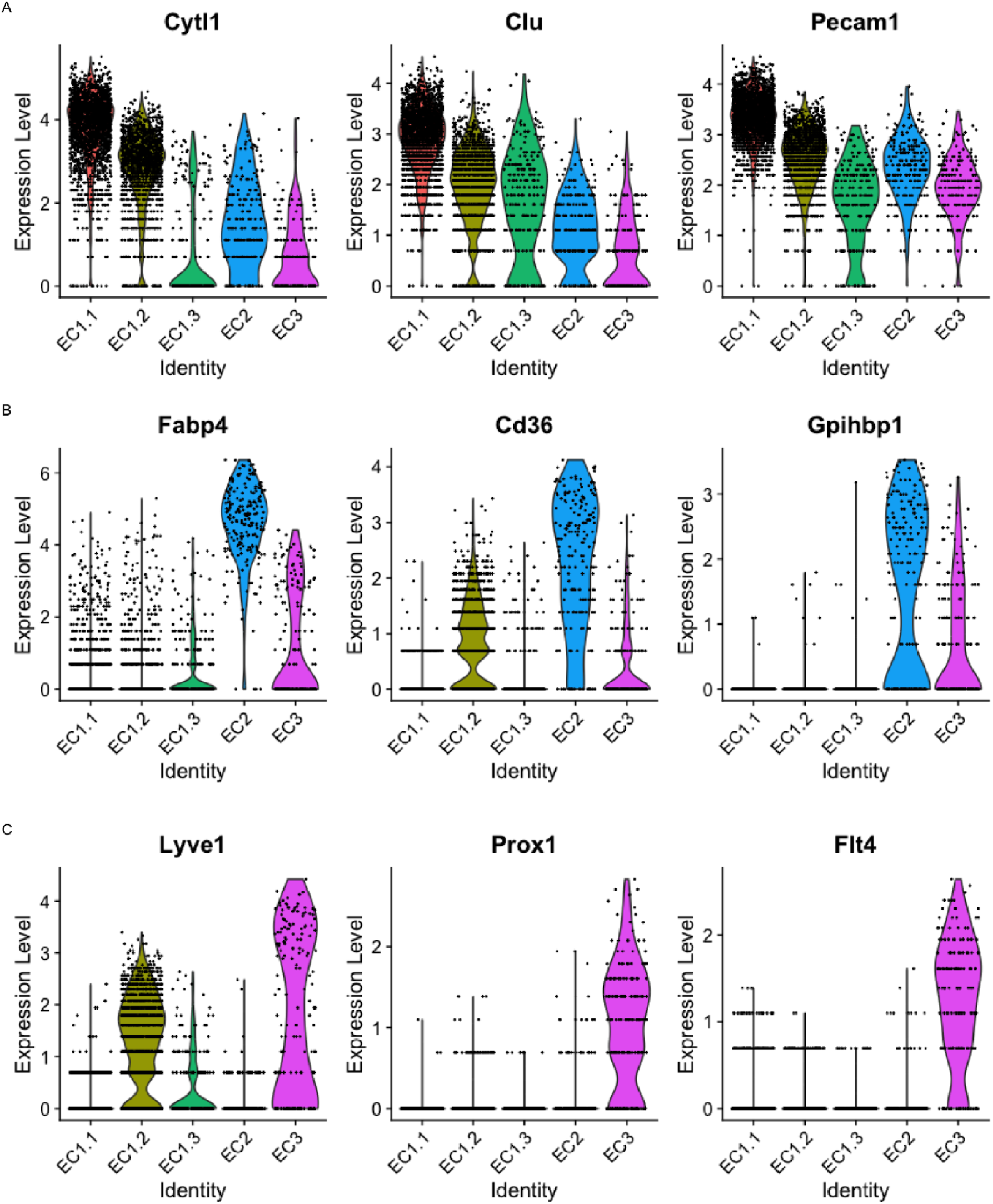
Expression of EC subcluster markers. Expression levels of published cluster markers for (**A**) EC1, (**B**) EC2, (**C**) EC3 (PMID: 31146585).

**Fig. S7.**
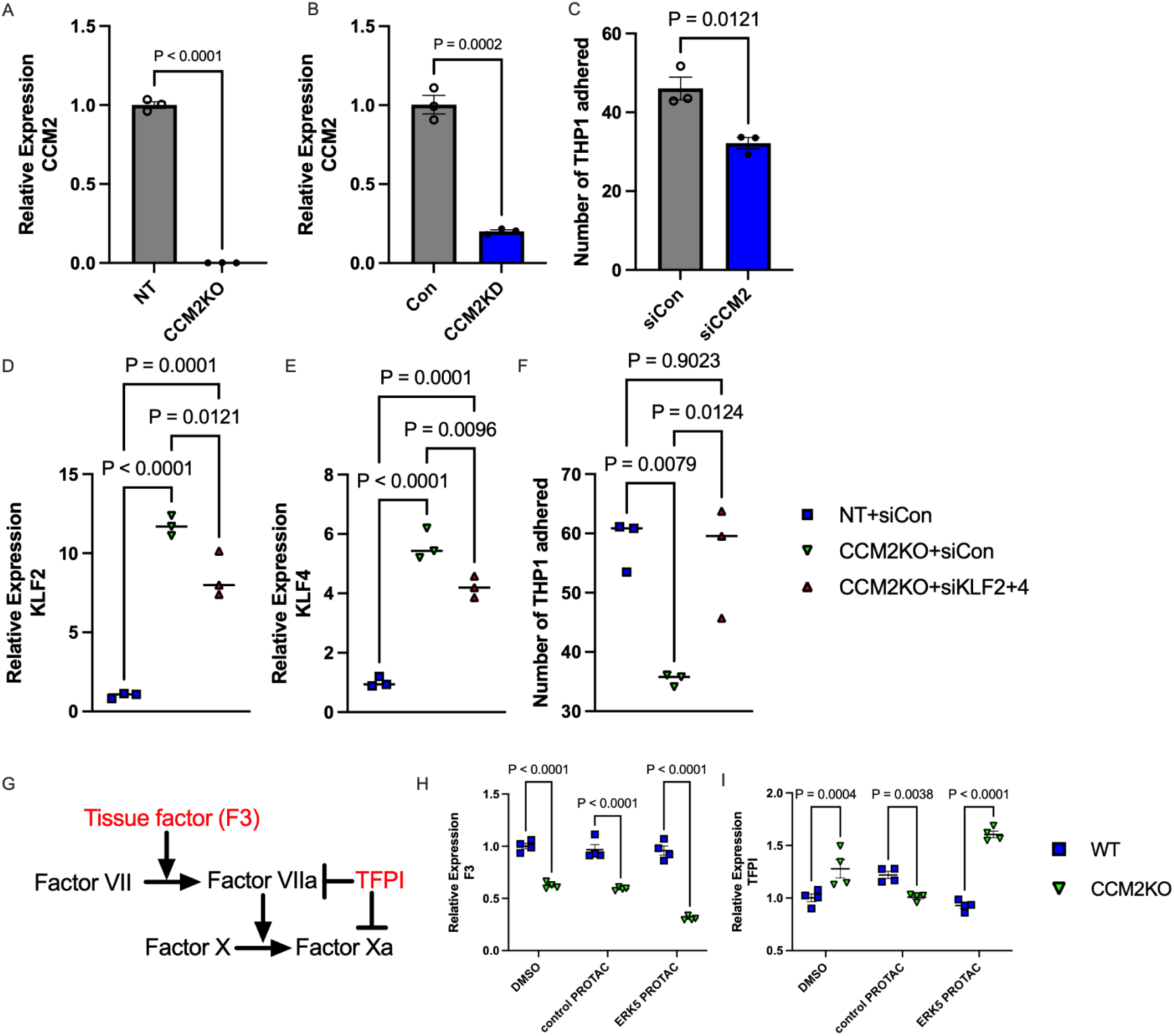
Validation of *CCM2* loss-of-function and *KLF2/4*-dependent regulation of monocyte adhesion and procoagulant responses in teloHAECs. (A-B) *CCM2* mRNA levels in teloHAECs with CRISPR-Cas9-mediated *CCM2* knockout (*CCM2*KO, **A**) or CRISPRi-mediated *CCM2* knockdown (*CCM2*KD, **B**). (**C**) Monocyte adhesion assay in teloHAECs with siRNA-mediated *CCM2* knockdown (siCCM2) or control nontargeting siRNA (siCon). (**D-E**) Relative mRNA levels of *KLF2* (**D**) and *KLF4* (**E**) in NT or *CCM2*KO teloHAECs treated with or without *KLF2/4* siRNA (siKLF2+4) or control (siCon). (**F**) Monocyte adhesion assay in NT or *CCM2*KO teloHAECs treated with *KLF2/4* or control siRNA. (**G**) Schematic of the Factor X activation pathway, highlighting the roles of *F3* and *TFPI*. (**H-I**) Expression of *F3* (**H**) and *TFPI* (**I**) in NT (WT) or *CCM2*KO teloHAECs treated without PROTAC (DMSO) or with the ERK5 or negative control PROTAC. Bars in (A)-(C): SEM. Each data point represents one biorep, calculated as the average of technical replicates. For (C) & (F) dots are the number of THP-1 cells per 1000 sorted TeloHAEC cells from separate wells.

**Fig. S8.**
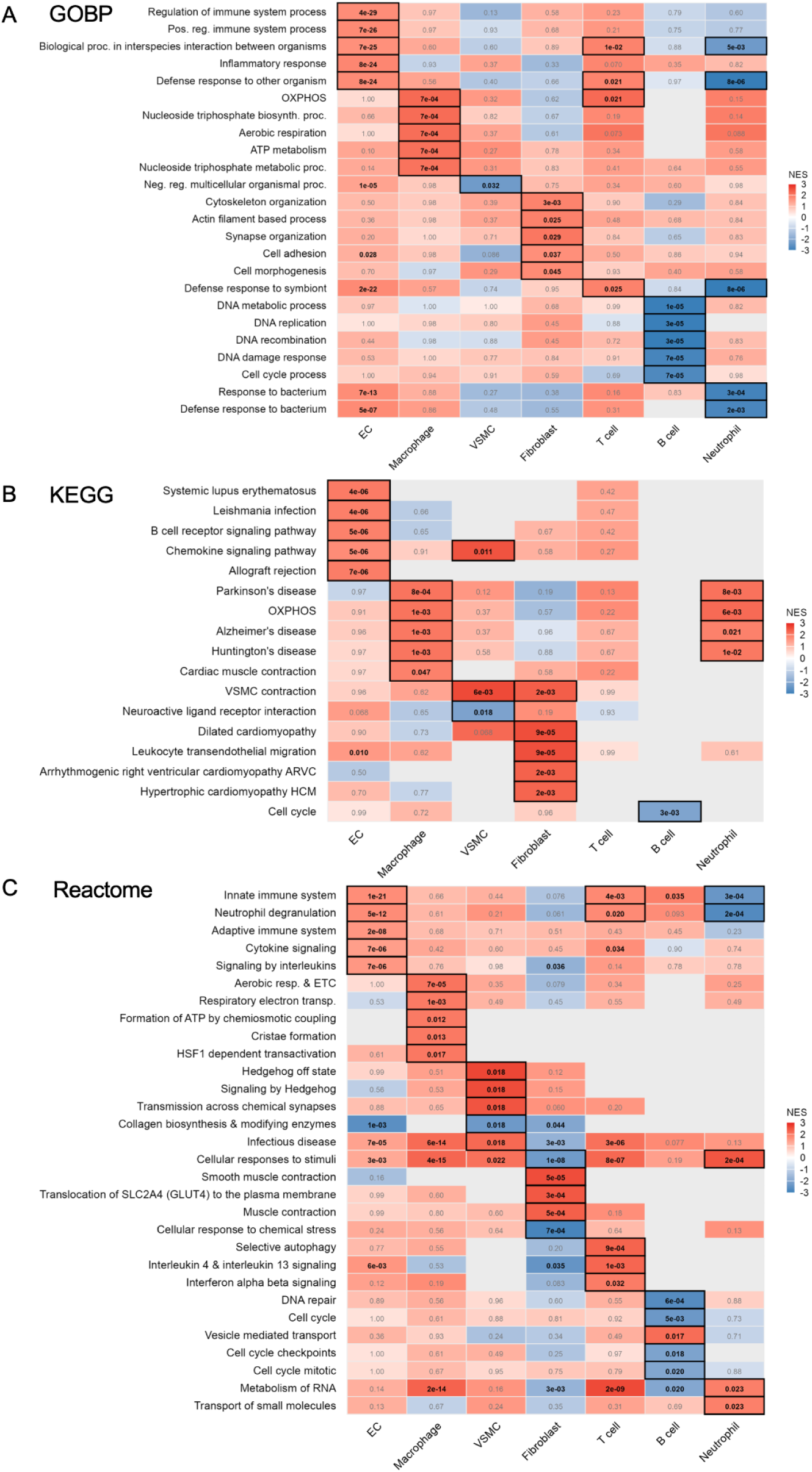
GSEA analysis with GOBP/KEGG/Reactome terms at Week 12. (**A**) Top 5 enriched pathways using GOBP terms, ranked strictly by -log_10_FDR in each aortic cell type *Ccm2*^+/-^ - *Ccm2*^+/+^. (**B**) As in (A), using KEGG terms. (**C**) As in (A), but using Reactome terms. NES, normalized enrichment score. Numbers in boxes, enrichment FDR.

**Fig. S9.**
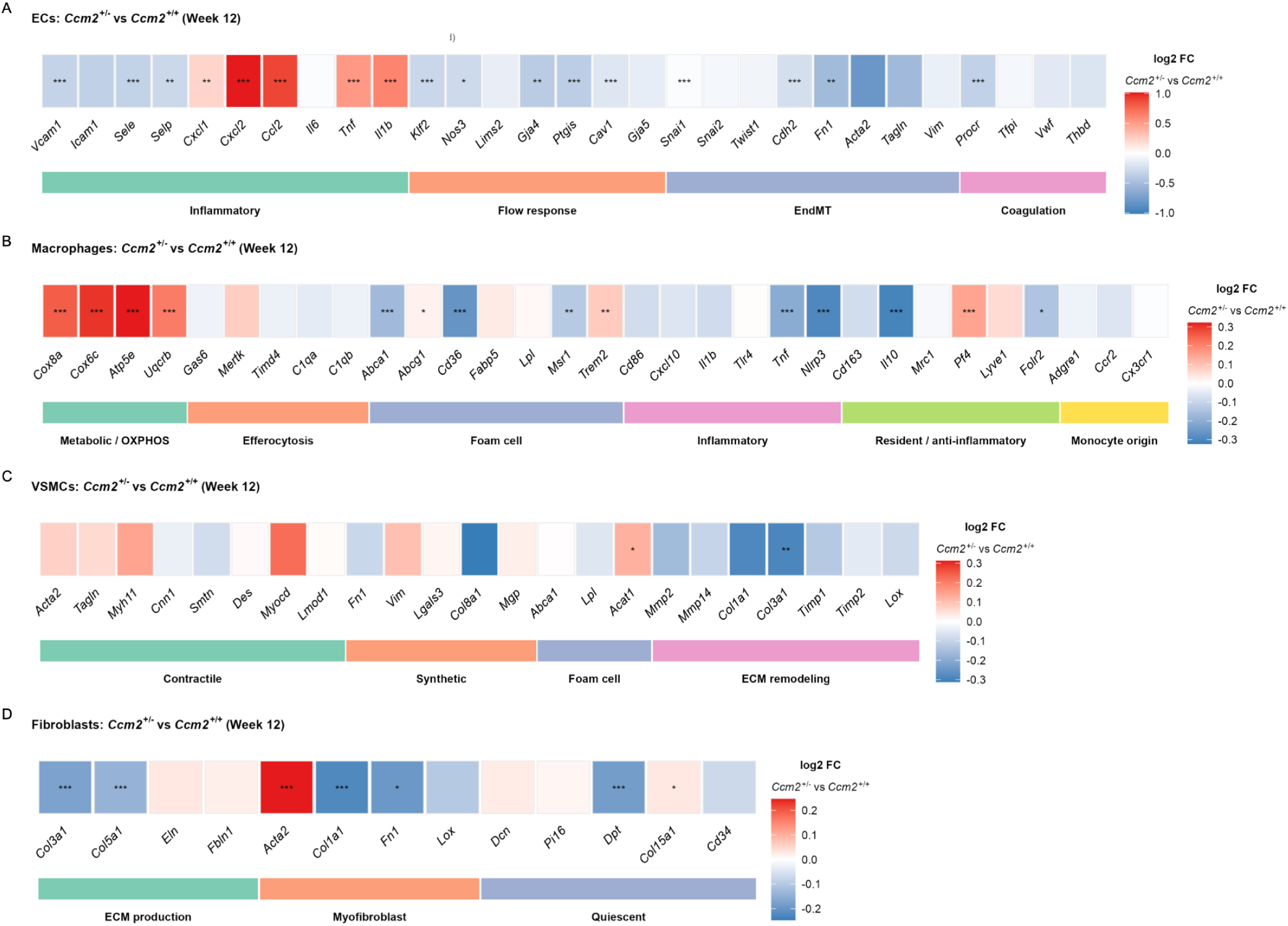
Differential expression of genes of interest in aortic cell types at Week 12. Log_2_FC of key genes in (**A**) ECs, (**B**) Macrophages, (**C**) VSMCs, and (**D**) Fibroblasts comparing *Ccm2*^+/-^ to *Ccm2*^+/+^ at Week 12. *FDR < 0.05, **FDR < 0.01, ***FDR < 0.001. Gene selection as per fig. S4.

**Fig. S10.**
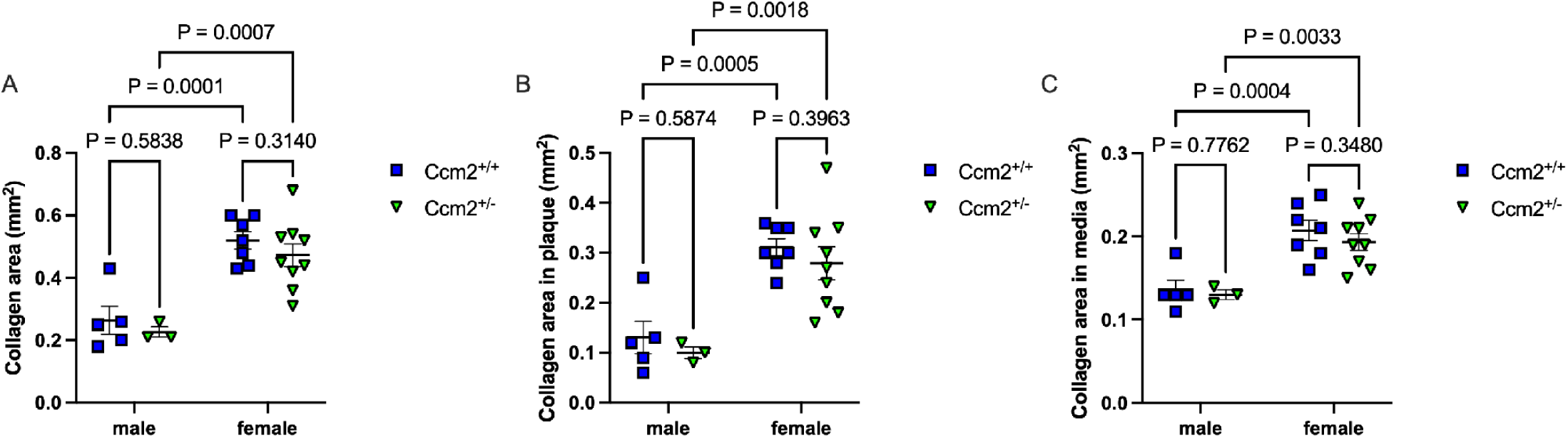
Collagen content in aortic roots at Week 12. Collagen content was quantified as blue pixel area in Masson’s Trichrome-stained sections. (**A**) Collagen area in the total aortic root of male and female *Ccm2*^+/+^ and *Ccm2*^+/-^ mice. (**B**) Collagen area restricted to atherosclerotic plaques. (**C**) Collagen area restricted to the tunica media. Data are mean ± SEM. Statistical tests: Two-way ANOVA with Šídák’s multiple comparisons test.

**Fig. S11.**
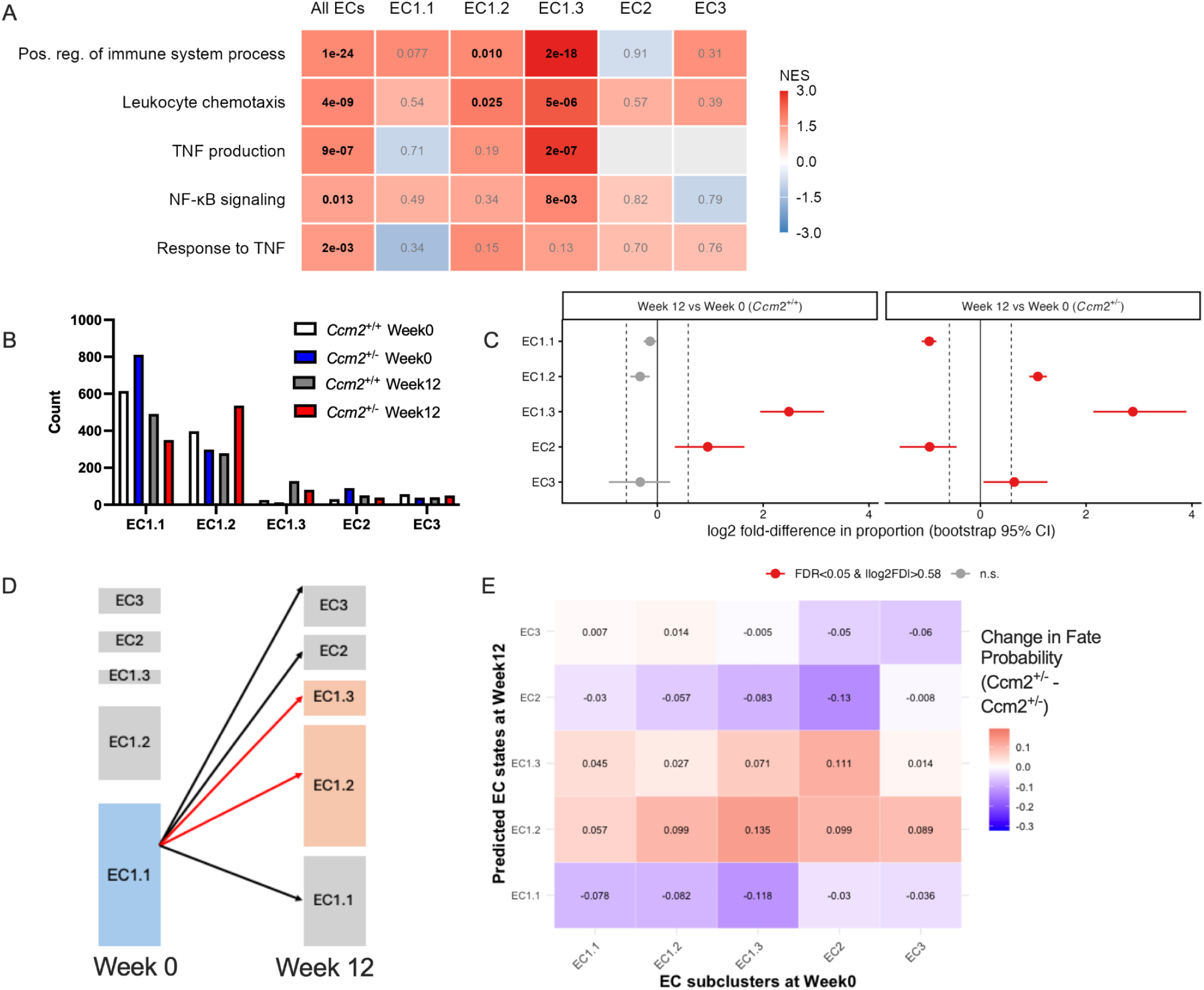
EC subcluster dynamics and cell state transitions in *Ccm2*^+/-^ aortas at Week 12. (**A**) Normalized enrichment scores (NES) for GOBP pathways across all ECs and individual EC subclusters. (**B**) EC subcluster cell counts in *Ccm2*^+/+^ and *Ccm2*^+/-^ aortas at Week 0 and Week 12. (**C**) The changes between EC subcluster frequencies in *Ccm2*^+/+^ or *Ccm2*^+/-^ aortas from week 0 to week 12. Red dots were called as significantly different by scProportion (FDR<0.05 and absolute fold change >1.5), but should be considered with the caveat that aortas in the same group were pooled before scRNA-seq. (**D**) Schematic of fate probability calculation for EC subclusters in WT cells. (**E**) Predicted change in cell fate probability (*Ccm2*^+/-^ - *Ccm2*^+/+^) of EC subclusters from Week 0 to Week 12.

**Fig. S12.**
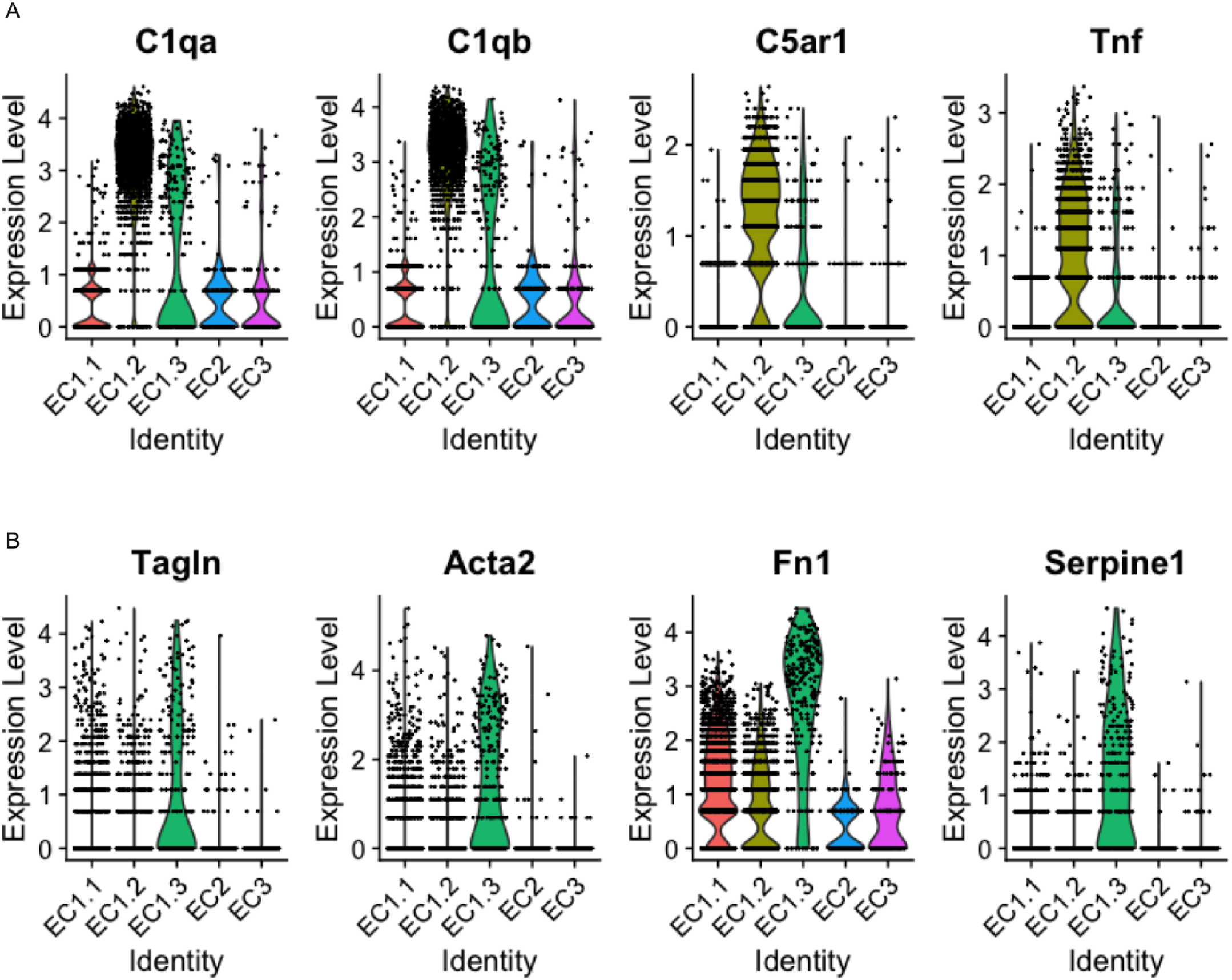
Validation of EC subcluster identity. (**A**) Expression of immune cell-like markers (*C1qa*, *C1qb*, *C5ar1*, *Tnf*) across EC subclusters (EC1.1, EC1.2, EC1.3, EC2, and EC3). (**B**) Expression of endothelial-to-mesenchymal transition (EndMT) markers (*Tagln*, *Acta2*, *Fn1*, *Serpine1*) across EC subclusters.

**Fig. S13.**
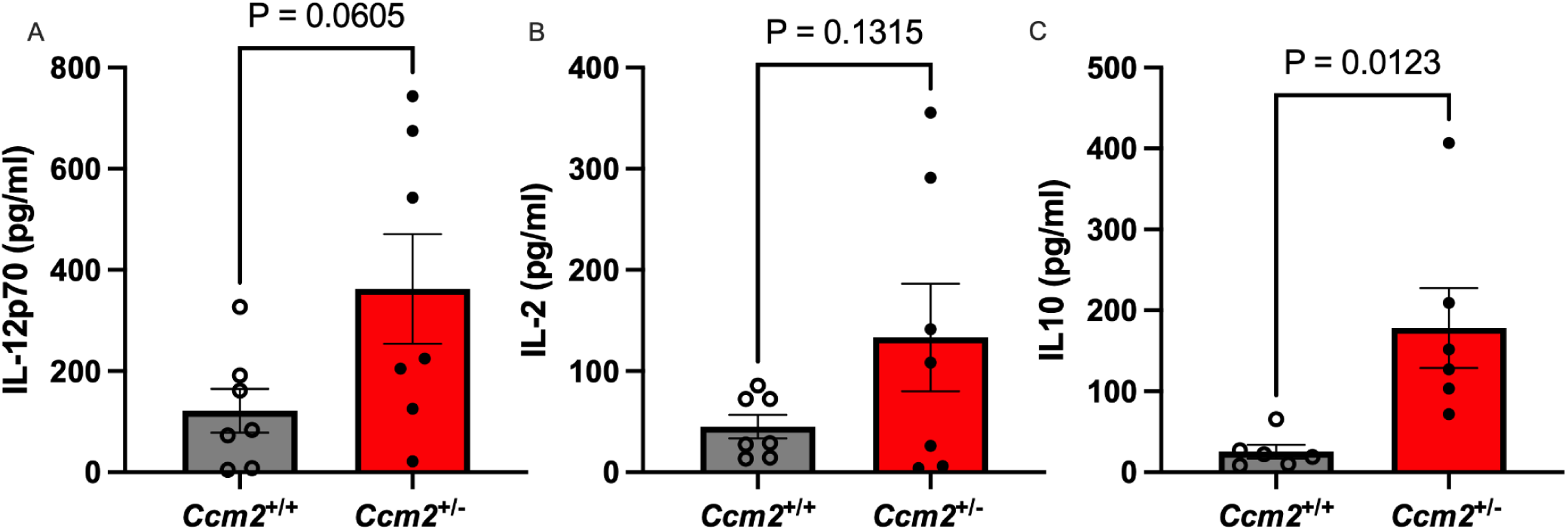
Plasma cytokine levels in mice at Week 12. Plasma levels of (**A**) IL-12p70, (**B**) IL-2, and (**C**) IL-10 in *Ccm2*^+/+^ and *Ccm2*^+/-^ mice (n = 5-7, male and female).

**Fig. S14.**
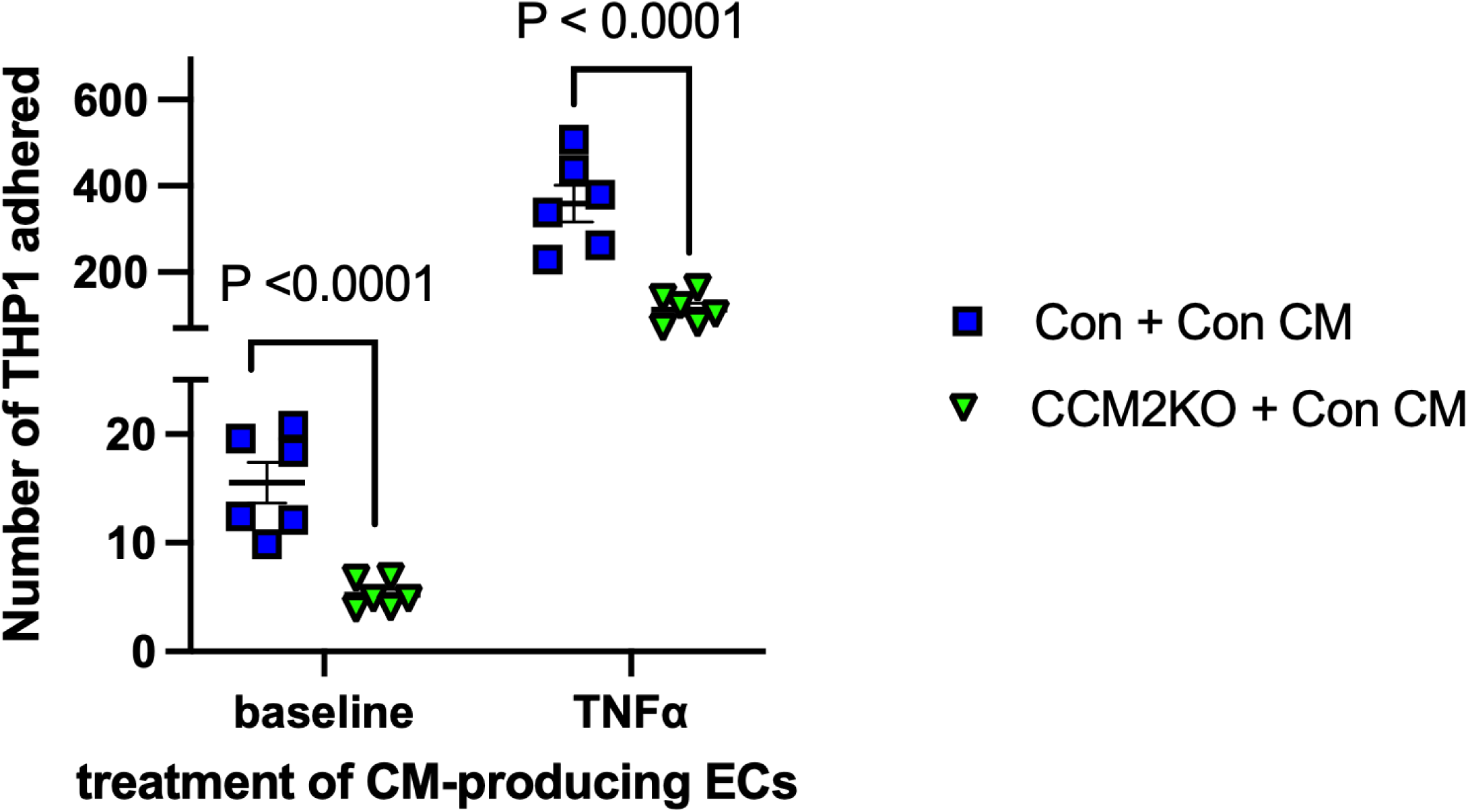
Reduced leukocyte adhesion in CCM2KO teloHAECs under conditioned media treatment. Monocyte adhesion to control (“Con”, teloHAECs transfected with non-targeting guides) or CCM2KO teloHAECs incubated with conditioned media collected from control teloHAECs under baseline or TNFα-stimulated conditions. Dots are the number of THP-1 cells per 1000 sorted TeloHAEC cells from separate wells.

**Fig. S15.**
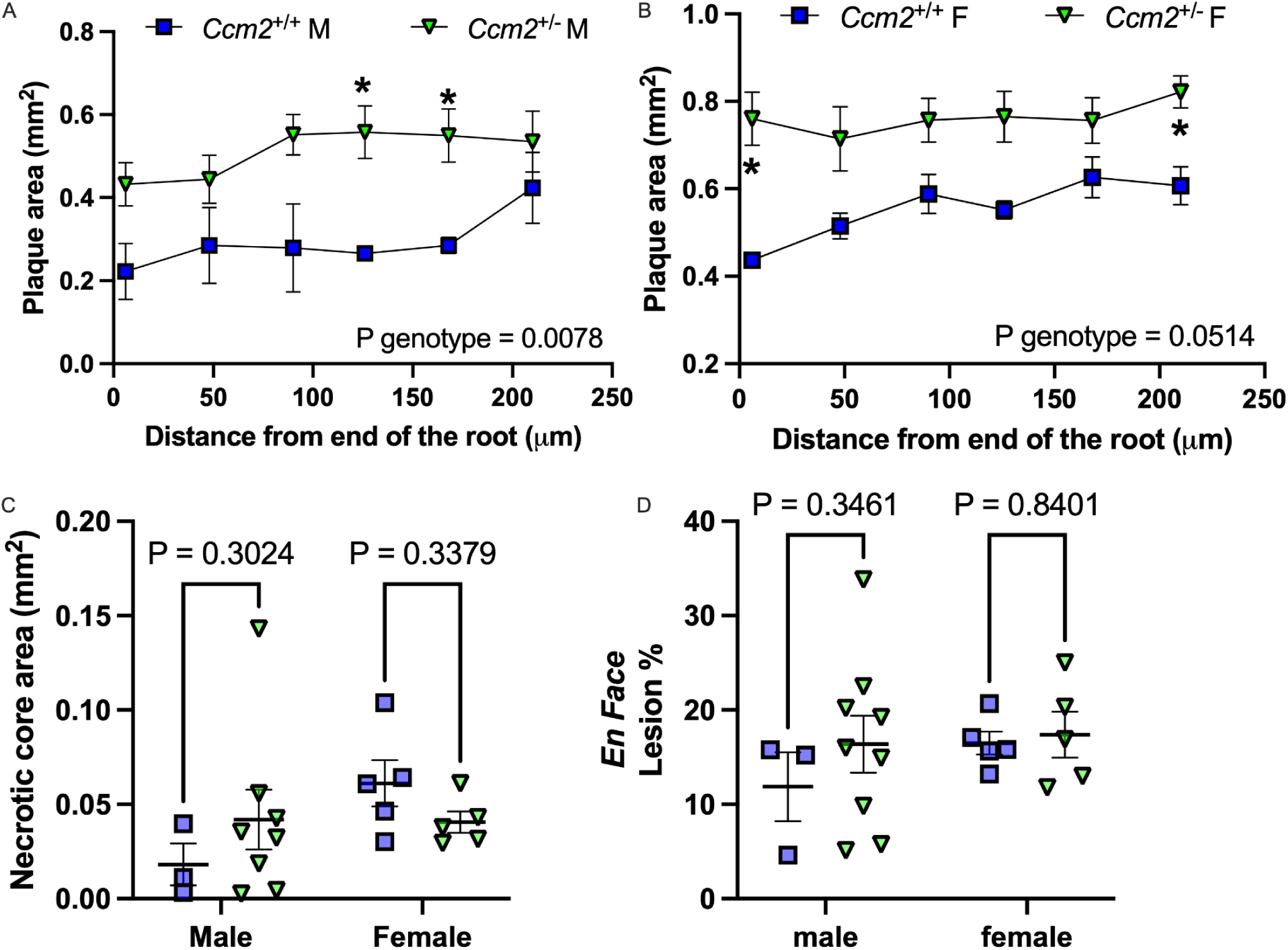
Atherosclerosis burden in mice with 20 weeks of atherogenic diet. (**A**-**B**) Aortic root plaque area in male (**A**) and female (**B**) mice. (**C**) Aortic root necrotic core area. (**D**) *En face* lesion area separated by sex. Data are displayed in mean±SEM. Two-way ANOVA with repeated measurements (A-B). Two-way ANOVA with Šídák’s multiple comparisons test (C,D).

**Fig. S16.**
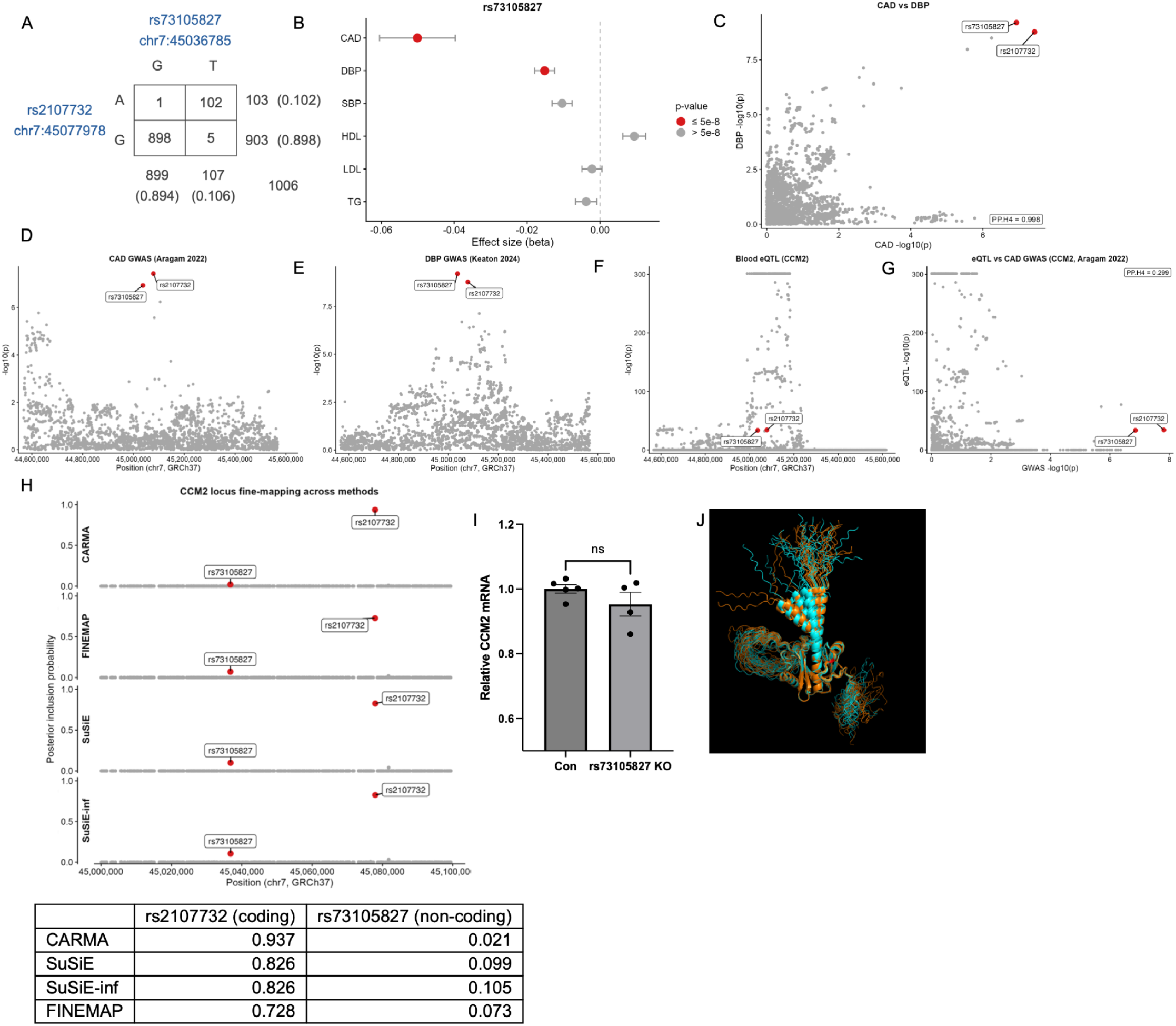
Variant prioritization of the *CCM2* locus at Chr7p13. (**A**) Haplotype frequency table for the top two variants in the *CCM2* locus. The rs2107732 (A) allele is in linkage disequilibrium with the rs73105827 (T) allele (R² = 0.9376, *P* < 0.0001). (**B**) PheWAS of non-coding variant rs73105827 demonstrating associations with decreased risk of CAD and DBP. (**C**) Colocalization analysis demonstrating shared signals for rs2107732 and rs73105827 in CAD and DBP GWAS. (**D**) Manhattan plot of CAD GWAS signals within a 1 Mb window of *CCM2*. (**E**) Manhattan plot of DBP GWAS signals within a 1 Mb window of *CCM2*. (**F**) Whole blood eQTL signals within a 1 Mb window of *CCM2*. (**G**) Colocalization analysis of CAD GWAS and blood eQTL. (**H**) Statistical fine-mapping of the *CCM2* locus using CARMA, FINEMAP, and SuSiE algorithms. PIP scores for the coding variant rs2107732 and non-coding variant rs73105827 are summarized in the table. (**I**) *CCM2* mRNA levels in teloHAECs with CRISPR-mediated truncation of 116 bp surrounding the non-coding variant rs73105827 (n=3). (**J**) AlphaFold simulation of the protein structure of WT CCM2 (orange) and V53I CCM2 (cyan).

**Fig. S17.**
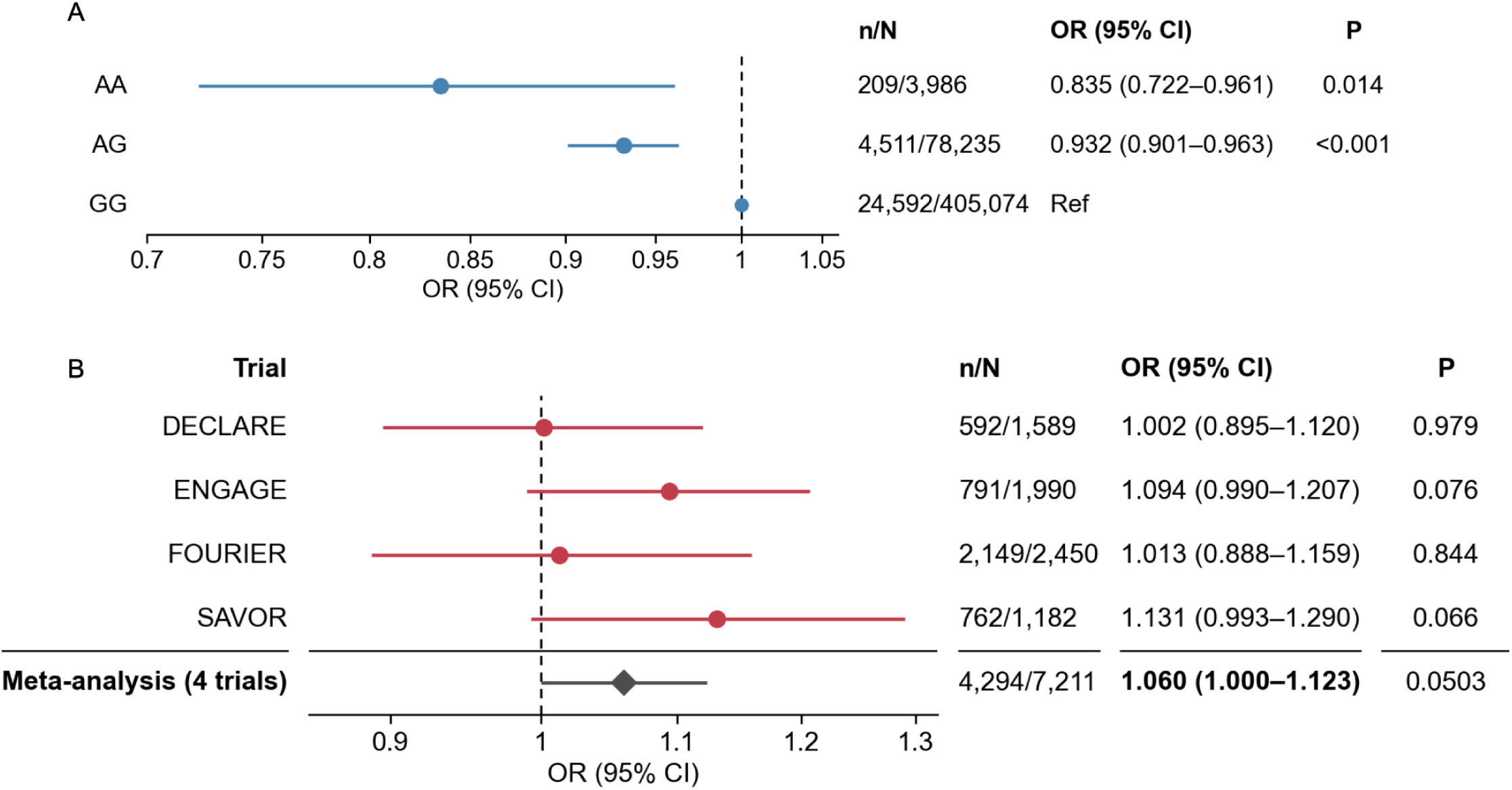
Association of *CCM2* coding variant V53I (rs2107732) with CAD risk across UK Biobank and TIMI clinical trial cohorts. (**A**) Association of rs2107732 genotype with combined and incident CAD in the UK Biobank. Data are displayed as odds ratio (OR) with 95% confidence interval (CI). G, major allele; A, minor allele. Models are adjusted for age, sex, and the first five genetic principal components. (**B**) Association of rs2107732 with CAD risk across individual TIMI trials. Meta-analysis of the four trials amenable to trial-specific analysis is shown (diamond). DECLARE: OR 1.002, 95% CI 0.895-1.120, P = 0.979; ENGAGE: OR 1.094, 95% CI 0.990-1.207, P = 0.076; FOURIER: OR 1.013, 95% CI 0.888-1.159, P = 0.844; SAVOR: OR 1.131, 95% CI 0.993-1.290, P = 0.066. SOLID-TIMI 52 and PEGASUS-TIMI 54 were excluded due to the lack of participants without an established ASCVD.

